# SARS-CoV-2 infection elucidates unique features of pregnancy-specific immunity

**DOI:** 10.1101/2024.02.05.24301794

**Authors:** Dong Sun Oh, Eunha Kim, Guangqing Lu, Rachelly Normand, Lydia L. Shook, Amanda Lyall, Olyvia Jasset, Stepan Demidkin, Emily Gilbert, Joon Kim, Babatunde Akinwunmi, Jessica Tantivit, Alice Tirard, Benjamin Y. Arnold, Kamil Slowikowski, MGH COVID-19 Collection & Processing Team, Marcia B. Goldberg, Michael R. Filbin, Nir Hacohen, Long H. Nguyen, Andrew T. Chan, Xu G. Yu, Jonathan Z. Li, Lael Yonker, Alessio Fasano, Roy H. Perlis, Ofer Pasternak, Kathryn J. Gray, Gloria B. Choi, David A. Drew, Pritha Sen, Alexandra-Chloé Villani, Andrea G. Edlow, Jun R. Huh

## Abstract

Pregnancy is a risk factor for increased severity of SARS-CoV-2 and other respiratory infections. The mechanisms underlying this risk have not been well-established, partly due to a limited understanding of how pregnancy shapes immune responses. To gain insight into the role of pregnancy in modulating immune responses at steady state and upon perturbation, we collected peripheral blood mononuclear cells (PBMC), plasma, and stool from 226 women, including 152 pregnant individuals (n = 96 with SARS-CoV-2 infection and n = 56 healthy controls) and 74 non-pregnant women (n = 55 with SARS-CoV-2 and n = 19 healthy controls). We found that SARS-CoV-2 infection was associated with altered T cell responses in pregnant compared to non-pregnant women. Differences included a lower percentage of memory T cells, a distinct clonal expansion of CD4-expressing CD8^+^T cells, and the enhanced expression of T cell exhaustion markers, such as programmed cell death-1 (PD-1) and T cell immunoglobulin and mucin domain-3 (Tim-3), in pregnant women. We identified additional evidence of immune dysfunction in severely and critically ill pregnant women, including a lack of expected elevation in regulatory T cell (Treg) levels, diminished interferon responses, and profound suppression of monocyte function. Consistent with earlier data, we found maternal obesity was also associated with altered immune responses to SARS-CoV-2 infection, including enhanced production of inflammatory cytokines by T cells. Certain gut bacterial species were altered in pregnancy and upon SARS-CoV-2 infection in pregnant individuals compared to non-pregnant women. Shifts in cytokine and chemokine levels were also identified in the sera of pregnant individuals, most notably a robust increase of interleukin-27 (IL-27), a cytokine known to drive T cell exhaustion, in the pregnant uninfected control group compared to all non-pregnant groups. IL-27 levels were also significantly higher in uninfected pregnant controls compared to pregnant SARS-CoV-2-infected individuals. Using two different preclinical mouse models of inflammation-induced fetal demise and respiratory influenza viral infection, we found that enhanced IL-27 protects developing fetuses from maternal inflammation but renders adult female mice vulnerable to viral infection. These combined findings from human and murine studies reveal nuanced pregnancy-associated immune responses, suggesting mechanisms underlying the increased susceptibility of pregnant individuals to viral respiratory infections.

## Introduction

Coronavirus disease 2019 (COVID-19) is a respiratory infectious disease caused by severe acute respiratory syndrome coronavirus 2 (SARS-CoV-2)^1–3^. The COVID-19 global pandemic has resulted in hundreds of millions of infections, including over two hundred thousand pregnant women in the United States alone^4^. While many studies have characterized immune responses to COVID-19 that are associated with disease progression in non-pregnant populations ^5–9^, substantially less is known about immune responses to COVID-19 in pregnancy^10–15^, despite the clinical observation that pregnant individuals are at increased risk for severe COVID-19-associated morbidity and mortality^16^. Pregnant individuals with COVID-19 are significantly more likely than age-matched non-pregnant individuals to require an intensive care unit (ICU) admission, invasive ventilation, and extracorporeal membrane oxygenation (ECMO), and they are more likely to die^17–19^. COVID-19 has also been associated with pregnancy-specific complications, including higher rates of preterm birth, postpartum hemorrhage, hypertensive disorders of pregnancy, and thrombotic events, resulting in increased maternal morbidity and mortality, as well as neonatal complications^20–22^. Furthermore, earlier studies suggested that hyperactivation of the maternal immune system might put fetuses at risk for adverse neurodevelopmental or metabolic programming^23–25^. Therefore, understanding the characteristics of pregnancy-specific viral immune responses is critical to developing effective treatments for pregnant individuals and to reducing pregnancy-associated severe morbidity, mortality, and other complications, for SARS-CoV-2 specifically and more broadly, for other viruses that may impact pregnant individuals and their offspring.

In severe or critical COVID-19, non-pregnant patients often exhibit aberrant activation of innate and adaptive immune responses^9,26^, leading to a profound functional alteration of immune cells^27,28^. For example, aggravated inflammatory responses accompanied by a massive increase in inflammatory cytokines are one of the key features of COVID-19, which is also closely correlated with disease severity^29–34^. An increase in regulatory T cell (Treg) levels has been observed^35,36^, presumably as a compensatory mechanism to dampen immune responses. Additionally, an exhausted T cell phenotype with upregulated inhibitory receptor expression has been reported, particularly in those with severe or critical disease^37^.

Pregnancy poses unique challenges to the maternal immune system, which is tasked with tolerating the fetus while simultaneously protecting pregnant individuals and the developing fetus from invading pathogens. Altered cytokine expression has been reported in pregnant individuals with COVID-19, relative to uninfected pregnant women^20–22,38^. Studies of pregnancy-associated immune changes in COVID-19, however, have reported varied results^39^. For example, while some studies reported altered T cell activation in SARS-CoV-2-infected pregnant women compared to non-pregnant individuals^40,41^, others indicated comparable responses^11,42^. In addition, studies have reported conflicting results regarding augmented versus downregulated function of circulating monocytes in pregnant women with SARS-CoV-2 infection^41,43,44^. These seemingly disparate findings might be attributed to heterogeneity and imprecise phenotyping of pregnant women’s clinical presentation within and across studies; some of these studies did not consider or did not have sufficient sample size and patient diversity to consider key clinical features, such as SARS-CoV-2 disease severity, interval since infection, and presence of comorbidities, such as obesity, that may influence immune response. In addition, the majority of prior studies have a relatively narrow focus on specific features of the immune response, rather than capturing comprehensive phenotyping of diverse immune cell populations, as well as the baseline and stimulated T cell, monocyte, and peripheral blood cytokine responses. Finally, few studies have comprehensively profiled the pregnancy-specific SARS-CoV-2 response, including critical comparator groups including with pregnant uninfected controls, non-pregnant controls, and non-pregnant individuals with COVID-19.

To address these critical gaps, we comprehensively assessed the differences between host immune responses to SARS-CoV-2 in pregnant women and non-pregnant women, including the effects of disease severity, phase of illness (acute versus convalescent), and obesity, the most common pregnancy comorbidity impacting the maternal host immune response. We also considered fetal sex in our analyses, which has been demonstrated to influence maternal and placental immune responses to SARS-CoV-2^12^. Using flow cytometry, single-cell multiomics, *ex vivo* immune cell stimulation, and unbiased immune correlative analyses, we performed comparative analyses aimed at defining pregnancy-associated immune changes during and following SARS-CoV-2 infection. We collected peripheral blood mononuclear cells (PBMCs), plasma, and stool from unvaccinated pregnant individuals with or without first-time SARS-CoV-2 infection, and non-pregnant women with or without first-time SARS-CoV-2 infection (**Table S1** and **Table S2**) as controls.

We found that CD8^+^ T cells in pregnant patients with severe/critical COVID-19 expressed elevated T cell exhaustion markers, such as programmed cell death-1 (PD-1) and T cell immunoglobulin and mucin domain-3 (Tim-3), compared to non-pregnant or convalescent patients with COVID-19. We also identified decreased anti-inflammatory Treg percentages in pregnant patients with severe/critical COVID-19 relative to those in non-pregnant women with severe/critical COVID-19. Further, we observed significantly down-regulated interferon-stimulated gene (ISG) responses in pregnant individuals with COVID-19 compared to non-pregnant COVID-19 counterparts. We identified altered T cell clonal expansion in pregnant patients with COVID-19, with these patients exhibiting a non-canonical, clonal expansion of CD4-expressing CD8^+^ T cells, compared to non-pregnant patients with COVID-19 who had expansion of a conventional cytotoxic CD8^+^ T cell subset, suggesting altered virus-mediated T cell responses in pregnancy. Relative to pregnant controls, COVID-19 infection in pregnancy was associated with suppressed monocyte function, manifested by decreased human leukocyte antigen (HLA)-DR protein levels and diminished cytokine and chemokine production. We also demonstrate that CD4^+^ and CD8^+^ T cells from obese pregnant women with COVID-19 produced increased inflammatory cytokine levels when stimulated *in vitro* compared to non-obese pregnant individuals with COVID-19. Intriguingly, we found pregnancy was associated with a significantly increased level of circulating interleukin (IL)-27, a cytokine involved in T cell exhaustion^45,46^. Using preclinical models, we demonstrated that augmented IL-27 resulted in more severe lung damage and increased mortality upon induction of respiratory viral infection, while protecting fetuses from maternal inflammation during pregnancy. Lastly, we found that pregnant patients with COVID-19 have distinct gut bacterial species compared to pregnant uninfected individuals and non-pregnant patients with COVID-19, highlighting the potential contribution of the gut microbiome in pregnancy-specific immune responses.

Collectively, these findings highlight novel mechanisms by which pregnancy influences maternal host immune responses against viral infection. Our work also provides a mechanistic framework that enables a comprehensive understanding of the impact of SARS-CoV-2 infection in pregnancy on the short- and longer-term immunity of both mothers and offspring.

## Results

### Comparative immune cell analyses of non-pregnant versus pregnant women with COVID-19

To decipher the critical immunological drivers shaping the host immune responses in pregnant women infected with SARS-CoV-2, PBMCs, plasma, and stool samples were collected from 226 women, including 152 pregnant and 74 non-pregnant women (**Tables S1** and **S2**). During the height of the COVID-19 pandemic (April 2020-January 2021), samples were prospectively collected from pregnant, unvaccinated women (see Methods) who had mild, moderate, and severe/critical COVID-19 (severity determinations were categorized as per NIH criteria^47,48^), and from uninfected contemporaneously pregnant controls (SARS-CoV-2 PCR negative). Samples were collected during the period of acute illness (within 30 days of positive SARS-CoV-2 PCR test) and/or during the convalescent period (>30 days after positive PCR test, see Methods). PBMCs from non-pregnant women with COVID-19 (n = 55) and from uninfected non-pregnant controls (n = 19) of similar age group were analyzed as additional comparator groups. We performed 25 multicolor flow cytometry analyses to profile immune cell composition and intracellular cytokine expression across 156 PBMC specimens (**Figure 1A, Figures S1A** and **B, Tables S1–3**).

**Figure 1.**
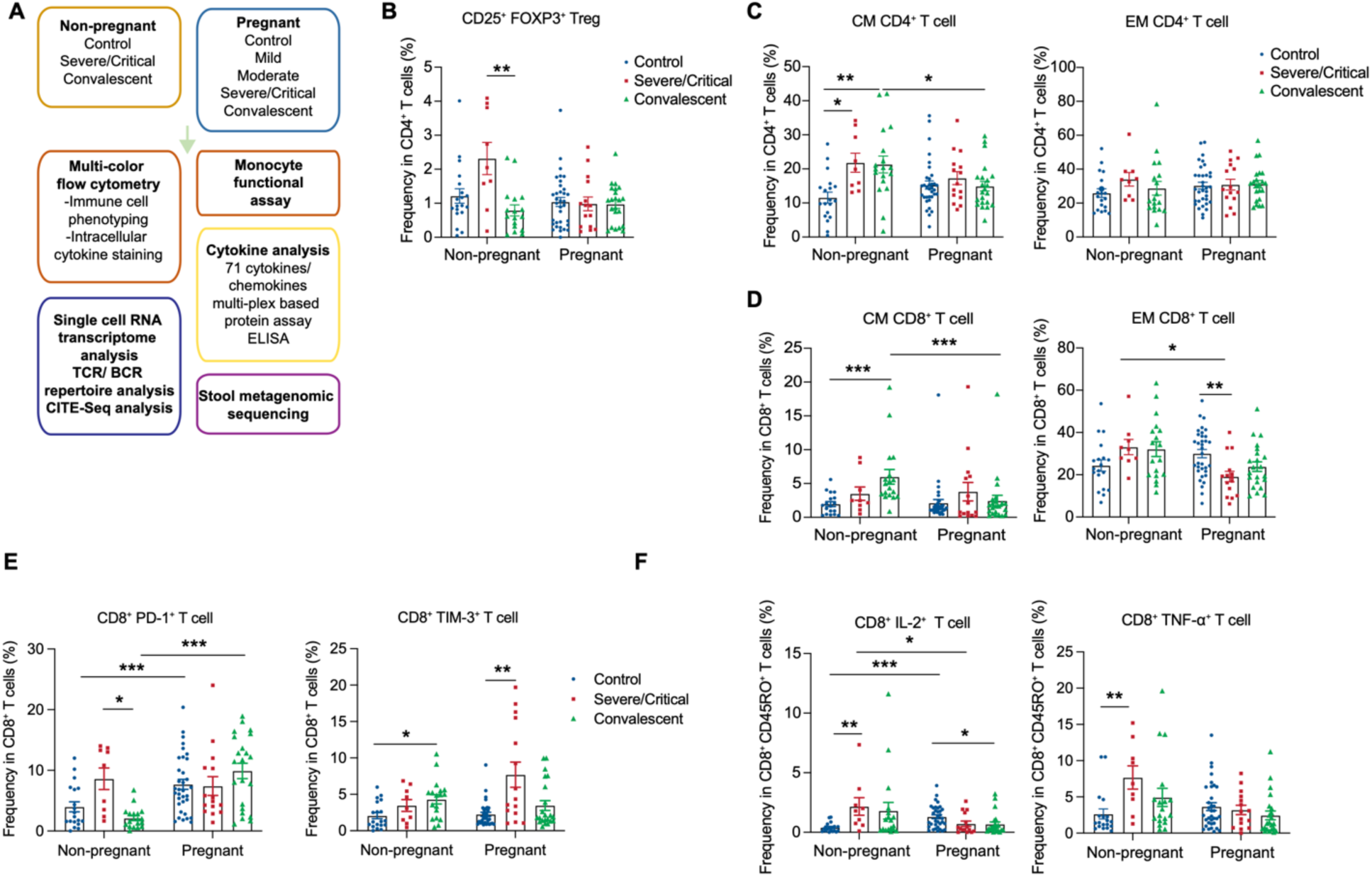
Comparative T cell analyses of pregnant versus non-pregnant patients with COVID-19. (**A**) A schematic of the analyses performed in this paper: non-pregnant versus pregnant women with or without COVID-19. (**B**) CD25^+^ FOXP3^+^ expressing CD4^+^ T cell frequencies in PBMCs. (**C** and **D**) Left panels show CM (CD45RO^+^ CCR7^+^) and right panels show EM (CD45RO^+^ CCR7^-^) frequencies in CD4^+^ T cells (CD19^-^ CD3^+^ CD4^+^) (**C**), and CD8^+^ (CD19^-^ CD3^+^ CD8^+^) T cells (**D**) in PBMCs. (**E** and **F**) Frequencies of CD8^+^ T cells expressing PD-1 and Tim-3 (**F**) and IL**-**2 and TNF-α. Healthy (Non-pregnant n = 19, Pregnant n = 34) or who had Severe/Critical COVID-19 (Non-pregnant n = 9, Pregnant n = 15) or convalescent COVID-19 (Non-pregnant n = 18, Pregnant n = 22). Data are shown as the mean ± SEM. *p < 0.05, **p < 0.01, ***p < 0.001.

Uncontrolled inflammatory responses are closely associated with poorer outcomes in COVID-19^49–54^. Tregs are often responsible for restraining excessive immune responses, and previous work has found that Treg frequencies were increased in severe COVID-19 patients^55^. Tregs also play critical roles in maintaining maternal-fetal tolerance^56^, and impairment of such tolerance leads to adverse pregnancy outcomes^57^. Thus, we first analyzed FOXP3^+^ CD25^+^ Treg frequencies among CD4^+^ T cells in PBMCs across six different groups analyzed consisting of non-pregnant and pregnant patients across three subgroups (control, severe, convalescent). Consistent with a previous report^55^, our data indicated Treg frequencies were increased in severe/critical non-pregnant COVID-19 cases, and were reduced to baseline, uninfected levels in the convalescent phase (**Figure 1B**). In contrast to these findings in non-pregnant women, in pregnant women, Treg frequencies did not increase with severe/critical COVID-19 (**Figure 1B**), suggesting that Tregs in pregnant patients with COVID-19 may not efficiently control inflammation. We next analyzed other immune cell frequencies among total live cells. Major populations such as T cell, B cell, monocyte, natural killer (NK), and natural killer T (NKT) cell levels among total live cells were comparable between non-pregnant and pregnant women regardless of their infection status or severity (**Figure S2A**). To investigate pregnancy-associated changes within a specific cell type, we analyzed cell frequencies at the subpopulation level.

Consistent with the percentage comparison among total live cells, we found T and B cell frequencies were comparable between non-pregnant and pregnant women with COVID-19 regardless of their disease severity (**Figure S2B**). An earlier report indicated that low-density neutrophil and eosinophil levels correlate with the severity of COVID-19 in non-pregnant individuals^58–60^. We found that neutrophil frequency within the CD3^-^CD56^-^CD66b^+^ population was significantly decreased in convalescent pregnant patients relative to convalescent non-pregnant individuals (**Figure S2B**). We also found a significant increase in NK cell frequencies in severe/critical COVID-19 in pregnancy, compared to pregnant controls. In contrast, there were no COVID-19–associated changes in NK cell frequencies in the non-pregnant group (**Figure S2B**). While NKT cell levels were increased in non-pregnant participants during COVID-19 convalescence, their levels were significantly decreased in the pregnant convalescent group compared to the severe/critical disease group. An increase in the γ8T cell percentage was previously reported in pregnant control women^61^, and a decrease in γ8T cell frequency has been noted in patients with COVID-19^62^. Consistent with prior findings, we observed that pregnant controls had significantly increased γ8T cell frequencies compared to uninfected non-pregnant women, and SARS-CoV-2 infection was associated with reduced γ8T cell fractions in both groups (**Figure S2C**). Unlike the non-pregnant groups, however, γ8T cell levels were not restored to the basal level in pregnant convalescent women (**Figure S2C**). Taken together, these data indicate that the frequency of various immune cells, particularly T cells, are differentially impacted by SARS-CoV-2 infection in pregnant compared to non-pregnant people.

### Pregnancy-dependent changes in T cell function

We next investigated whether pregnancy was associated with altered T cell activation and function in the setting of COVID-19. First, we analyzed major T cell subpopulations defined by CCR7 and CD45RO expression level measured by flow cytometry. In non-pregnant women with COVID-19 (n = 27), the percentage of the central memory (CM, CD45RO^+^, CCR7^+^) subset of CD4^+^ T cells was significantly increased in the severe/critical (n = 9) and convalescent COVID-19 subjects (n = 18) compared with uninfected non-pregnant controls (n = 19) (**Figure 1C**). CM CD8^+^ T cell levels were significantly increased in non-pregnant convalescent women compared with uninfected controls (**Figure 1D**). Such changes were not observed in the pregnant group. Pregnant convalescent individuals (n = 22) had significantly lower frequencies of CM CD4^+^ and CD8^+^ T cells relative to non-pregnant convalescent women (n = 18) (**Figures 1C** and **D**). Effector memory (EM, CD45RO^+^, CCR7^-^) CD8^+^ T cell percentages were lower in the pregnant patients with COVID-19 (n = 15) relative to both pregnant uninfected controls and non-pregnant women with COVID-19 (**Figure 1D**). These data indicate that the EM CD8^+^ T cell response, one of the most important T cell subsets for antiviral immune responses^63,64^, was likely impaired in pregnant women with COVID-19. Furthermore, reduced percentages of the CM CD8^+^ T cells in the pregnant convalescent group may indicate that the maintenance of CM CD8^+^ T cell function was impaired in pregnant women following COVID-19.

T cell exhaustion and associated functional impairment phenotypes have been previously reported in patients with COVID-19^26^ characterized by the upregulation of inhibitory receptors, such as PD-1 and Tim-3^65,66^. In addition, upregulation of PD-1 and Tim-3 in peripheral leukocytes has also been noted in normal pregnancy and is thought to be associated with maternal-fetal immune tolerance^66^. Consistent with these prior findings, PD-1 expression was increased in CD8^+^ T cells in the pregnant uninfected control (n = 34) and pregnant convalescent (n = 22) groups, compared with their non-pregnant control and convalescent counterparts (**Figure 1E**). SARS-CoV-2 infection was associated with increased T cell exhaustion in pregnant patients with severe/critical COVID-19 (n = 15), as indicated by significantly increased TIM-3 levels compared to pregnant uninfected controls (**Figure 1E**). These data indicate that CD8^+^ T cell function may be impaired in the pregnant group.

We further assessed the impact of pregnancy on T cell function by measuring the production of cytokines upon *in vitro* stimulation of PBMCs with phorbol 12-myristate 13-acetate (PMA) and ionomycin. We found that, unlike non-pregnant patients with COVID-19 (n = 27), IL-2 and tumor necrosis factor-alpha (TNF-α) production by CD45RO^+^CD8^+^ T cells in pregnant patients with COVID-19 (n = 37) were not increased compared to control groups (**Figure 1F**). Severe/critical COVID-19 induced increased production of IL-2, TNF-α, interferon (IFN)-γ, and granzyme B by CD45RO^+^CD4^+^ T cells in the non-pregnant group (**Figure S3A**). Such an increase was not observed in pregnant women with COVID-19. Production of IL-17A, however, was significantly increased in the setting of severe/critical COVID-19 in pregnant women (n = 15) only (**Figure S3A**). We also noted that γ8T cells from pregnant women with COVID-19 produced increased levels of IFN-γ compared to control pregnant women (**Figure S3B**). These data suggest that CD8^+^ T cell function may be impaired in pregnant women due to the enhanced expression of co-inhibitory molecules, which may serve as a contributing factor to predispose pregnant women to increased disease severity in the setting of COVID-19. Conversely, T helper (Th)17 cell function seems to be enhanced in the setting of viral infection in pregnancy, echoing earlier findings from inflammation-exposed pregnant rodent work^67–69^.

To better understand COVID-19 severity within the pregnant individuals (**Tables S1** and **S2**), we next compared T cell function within pregnant groups depending on COVID-19 severity. Compared to other groups, EM CD4^+^ and EM CD8^+^ T cell frequencies were altered in the moderate (n = 12) and severe/critical groups (n = 15), respectively (**Figures S4A** and **S4B**). Upon *in vitro* stimulation, CD4^+^ T cells isolated from the moderate group produced the highest amounts of IFN-γ, TNF-α, and granzyme B, compared to other groups, including the severe/critical group (**Figure S4C**). Such moderate group-associated robust cytokine production was not observed in IL-17A–producing CD4^+^ cells (**Figure S4D**). IFN-γ–producing CD8^+^ T cell level was also highest in the moderate group (**Figure S4E**). On the other hand, IFN-γ– and TNF-α–producing γδT cell levels were strongly induced in the mild, not moderate, COVID-19 group (**Figure S4F**).

Taken together, these data indicate that SARS-CoV-2 infection led to altered T cell function, which was more pronounced in particular cell types depending on disease severity (e.g. CD4^+^ and CD8^+^ T cells were less active in the severe/critical pregnant COVID-19 group compared to the moderate group). Moreover, our results are consistent with altered T cell function in pregnancy, particularly that of CD8^+^ cells, and suggest the enhanced expression of co-inhibitory markers may underlie T cells’ impaired activity.

### Pregnancy-dependent changes in monocyte function

We next assessed whether pregnancy and COVID-19 were associated with alterations in monocyte composition and function. Three major populations of monocytes can be defined by flow cytometry in blood based on the level of CD14 and CD16 expression: classical monocytes, CD14^+^CD16^-^; intermediate monocytes, CD14^+^CD16^+^; and non-classical monocytes, CD14^low^CD16^+70,71^. Classical monocyte levels were highest in pregnant convalescent women (n = 22) compared to other groups (**Figure 2A**). Levels of intermediate monocytes were increased in non-pregnant individuals with severe-critical COVID-19 (n = 9) relative to uninfected controls (n = 19) (**Figure 2A**); this increase was not observed in pregnant individuals. Finally, levels of non-classical monocyte frequencies were significantly higher in pregnant convalescent women (n = 22) compared to non-pregnant convalescent women (n = 18) (**Figure 2A**).

**Figure 2.**
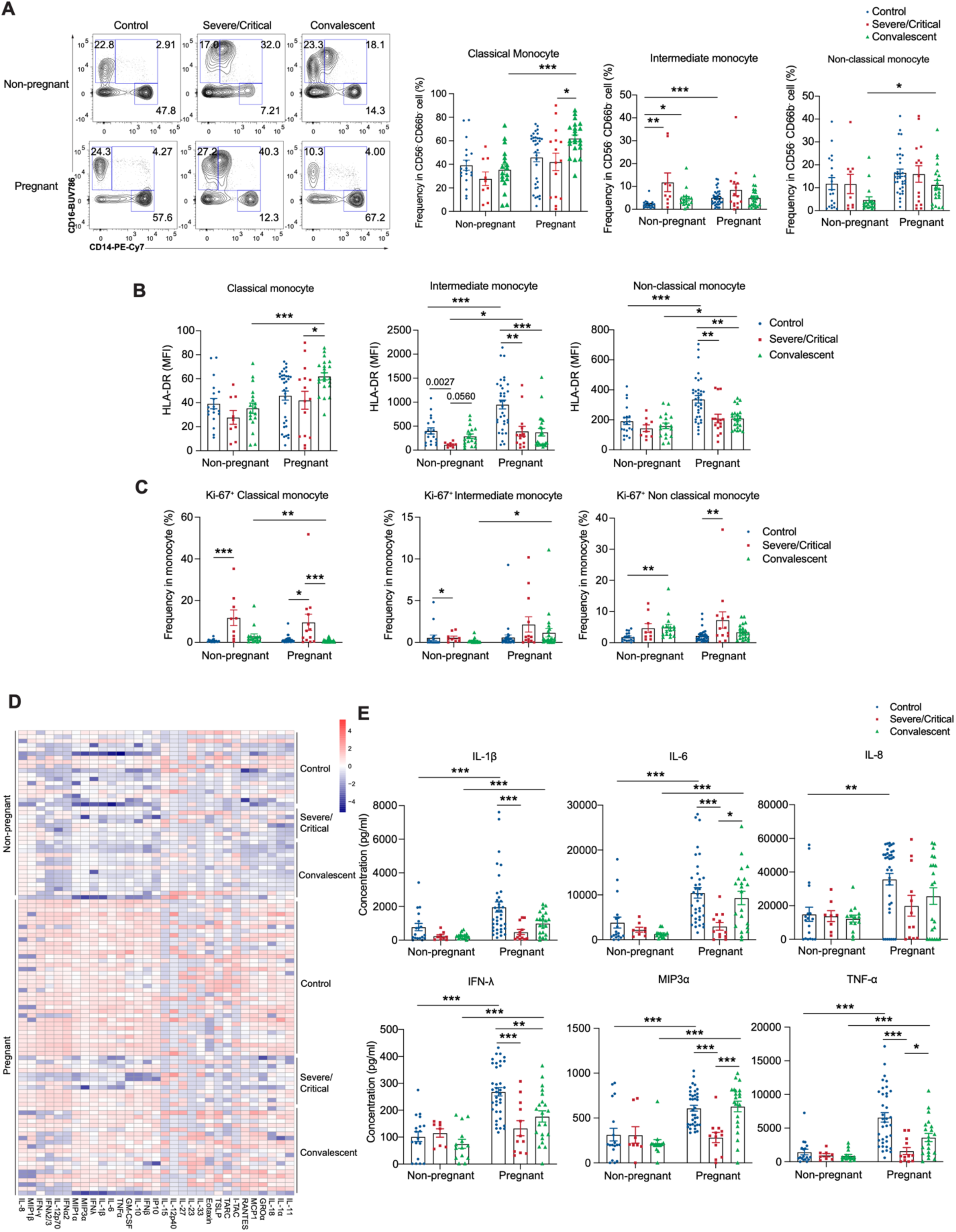
Pregnancy enhances monocyte function. (**A**) Representative FACS plots and quantifications of classical monocytes (CD14^+^), intermediate monocytes (CD14^+^CD16^+^), and non-classical monocytes (CD16^+^) as analyzed by flow cytometry from PBMCs. (**B**) Mean fluorescent intensities (MFI) of HLA-DR expression in classical, intermediate, and alternative monocytes. (**C**) Frequencies of Ki67, a proliferation marker, expressed in classical, intermediate, and alternative monocytes. (**D** and **E**) A heatmap presenting monocyte-secreted cytokines upon 1 ng/mL LPS stimulation (**D**) and representative cytokine levels in the supernatant as quantified by CBA analyses (**E**). Healthy (Non-pregnant n = 19, Pregnant n = 34) or who had Severe/Critical COVID-19 (Non-pregnant n = 9, Pregnant n = 15) or convalescent COVID-19 (Non-pregnant n = 18, Pregnant n = 22). Data are shown as the mean ± SEM. *p < 0.05, **p < 0.01, ***p < 0.001.

We further examined monocyte expression of human leukocyte antigen-DR (HLA-DR), an MHC class II surface receptor that presents antigens to T cells. Earlier studies of patients with severe COVID-19 identified reduced HLA-DR expression on monocytes as a marker for immune suppression and monocyte exhaustion^27,28,72–74^. In intermediate and non-classical monocytes, HLA-DR expression levels were higher at baseline in the pregnant uninfected controls (n = 34) compared to non-pregnant (n = 19) controls. HLA-DR expression was significantly reduced in the setting of severe-critical COVID-19 relative to uninfected controls, in both pregnant and non-pregnant groups (**Figure 2B**). Intriguingly, in pregnant individuals, HLA-DR expression was restored during convalescence in classical monocytes only, while it remained suppressed even in convalescence in intermediate and non-classical monocytes. Moreover, HLA-DR expression in both classical and non-classical monocytes was significantly higher in the pregnant convalescent group relative to non-pregnant convalescent individuals (**Figure 2B**). Taken together, these findings suggest that pregnant individuals may have heightened antigen-presenting monocyte capabilities compared to non-pregnant individuals, with antigen-presenting capacity of monocytes significantly reduced in the setting of severe or critical COVID-19. The observed reduction in HLA-DR expression likely reflected monocyte exhaustion rather than reduced monocyte proliferation, as staining with the active proliferation marker Ki67 revealed robust proliferation of classical and non-classical monocytes during the acute phase of COVID-19 in both non-pregnant and pregnant patients (**Figure 2C**). In aggregate, these data indicate that pregnancy itself alters the relative abundance of certain circulating monocyte subsets, and thus the monocyte exhaustion associated with severe-critical COVID-19 may have a differential impact on the antigen-presenting capabilities of pregnant individuals, and therefore their ability to activate specific T cell responses. To test monocyte function in pregnant individuals with and without COVID-19, we enriched monocytes from PBMCs using CD14 microbeads, followed by *ex vivo* stimulation with a low amount of lipopolysaccharide (LPS) (1 ng/ml; see Methods). For this stimulation assay, we selected the concentration of LPS level that did not trigger robust cytokine responses in CD14^+^ monocytes isolated from the non-pregnant uninfected controls (**Figure 2D**). CD14^+^ monocytes from pregnant uninfected individuals produced higher mean levels of many cytokines compared to CD14^+^ monocytes from all non-pregnant groups (**Figure 2D**). In the setting of COVID-19, however, particularly in severe/critical patients, cytokine production by monocytes from pregnant individuals was significantly dampened relative to the pregnant uninfected baseline and relative to convalescent pregnant individuals (**Figure 2E**). For example, IL-1β, IL-6, IFN-λ, CC chemokine ligand 20 (CCL20, MIP3a), and TNF-α levels were significantly reduced in pregnant patients with severe/critical COVID-19 relative to uninfected pregnant controls, and were comparable between the pregnant severe/critical COVID-19 patients and the non-pregnant groups (**Figure 2E**). In sharp contrast to non-pregnant individuals, pregnant convalescent patients’ monocytes regained the ability to produce high amounts of many cytokines (e.g., IL-1β, IL-6, MIP3a, and TNF-α) (**Figure 2E**). We then compared monocyte function among pregnant women with different disease severity. While monocytes from the convalescent group display enhanced MIP3a and IL-6 production compared to the severe/critical group (**Figure S5A**), SARS-CoV-2 infection in pregnancy led to a prolonged suppressive effect on monocyte function, as evidenced by significantly reduced production of IL-12p70, IFN-α, and IFN-γ (**Figure S5B**). Thus, monocytes from pregnant individuals demonstrated an enhanced ability to produce cytokines at baseline, which was robustly suppressed by severe/critical COVID-19 to a greater extent than monocytes from non-pregnant counterparts. Of note, monocytes from pregnant individuals recovered their cytokine-producing function in convalescence, while such recovery was not observed in monocytes from non-pregnant individuals.

### Immune responses associated with pregnancy defined by single-cell RNA sequencing (scRNA-seq) of PBMCs from pregnant women with COVID-19

To gain an unbiased understanding of the differences in host immune responses between pregnant and non-pregnant patients with COVID-19, we performed scRNA-seq with cellular indexing of transcriptomes and epitope sequencing (CITE-seq) on 26 PBMC samples, including those from pregnant patients with severe COVID-19 (n = 3), asymptomatic COVID-19 (n = 3) or no SARS-CoV-2 infection (n = 3); as well as age- and sex-matched controls from non-pregnant female patients with severe COVID-19 (n = 11), and asymptomatic COVID-19 (n = 6), see **Table S3**). We analyzed a total of 181,112 single cells which grouped into the following circulating immune cell lineages: T (CD4^+^ and CD8^+^) and NK cells (95,225 cells), mononuclear phagocytic (MNP) cells (66,719 cells), and B and plasma cells (19,168 cells, **Figures S6A–C**). There were no significant differences in the abundance of these main lineages between pregnant and non-pregnant patients with COVID-19 nor across COVID-19 severities (**Figures S6D** and **E**).

We then sub-clustered the CD8^+^ T, CD4^+^ T, B/plasma, and MNP cells independently to further define immune cell subpopulations within each lineage with greater granularity (**Figures S6F–I, S7A, S8A, S9A-B, see Methods**). First, we identified 16 distinct cell subsets within the CD8^+^ T cell lineages, including an NK/NKT cell subset (NK/NKT_1), cycling NK cell subset (NK_2), naïve CD8^+^ T cells (CD8T_4), two CD8^+^ T cell subsets expressing cytotoxic genes GZMK and GZMH (CD8T_1, CD8T_2), one CD8^+^ T cell subset with gene expression suggestive of metabolic activation (CD8T_6), and three CD161-expressing CD8^+^ T cell subsets (CD8T_9, 10, MAIT). We also identified a CD8^+^ T cell subset with high expression of *CD4* and low expression of *CD8A* and *CD8B* (CD8T_3) and a CD8+ T cell subset with a high expression of CXCR4 and TGFB1 (CD8T_7) (**Figure 3A**). The abundance of the cytotoxic population CD8T_2 expressing *PRF1*, *KLRD1*, and *GNLY* was significantly lower in pregnant versus non-pregnant subjects with COVID-19, as was the cell subset CD8T_9 – a *CD161*, *RORC*, *CXCR6*, and *GZMK* expressing CD8^+^ T cell population suggestive of a Tc17 phenotype. In contrast, in the context of COVID-19 infection, CD8T_10 – a population of CD161/KLRB1-expressing CD8^+^ T cells with high *CXCR4* and *TNFAIP3* expression – is significantly higher in pregnant patients; MAIT cells also showed a trend of higher abundance in pregnant patients (**Figure 3B** and **Figures S6J, K**). Collectively, these shifts in cell subset abundance suggest that pregnant individuals have an altered capacity to mount specific immune responses in the setting of SARS-CoV-2 infection compared to non-pregnant individuals, and this occurs across a spectrum of mechanisms, including less robust classic cytotoxic CD8^+^ T cell (CD8T_2) and mitigated Tc17 (CD8T_9) responses^15^. In parallel, our data demonstrates that pregnant individuals with COVID-19 may have higher levels of other specialized *KLRB1*-expressing CD8^+^ T cell responses (CD8T_10, MAIT, **Figure 3B**), which have been shown to play an important role in mediating antimicrobial defense while maintaining the immune tolerance state of pregnancy^75^.

**Figure 3.**
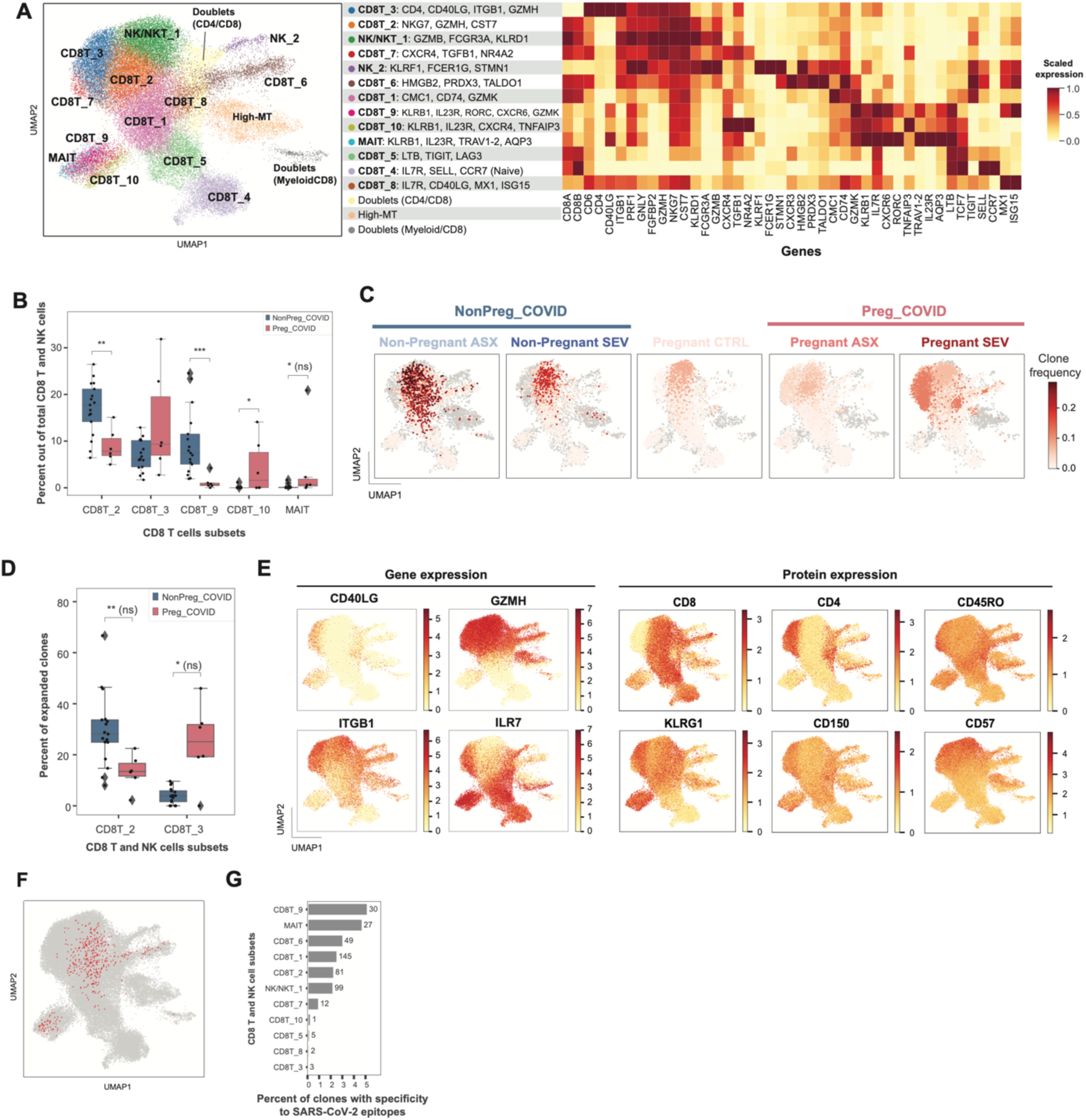
Changes in T cell clonal expansion and CD8^+^ T cell subset abundances in pregnant patients with COVID-19. (**A**) Uniform Manifold Approximation (UMAP) embedding of 50,140 CD8^+^ T and NK cells and their clustering to 13 cell subsets (left). Expression of main marker genes, scaled mean log(CPM) (right). (**B**) A boxplot illustrating the abundance of CD8^+^ T cell subsets out of all CD8^+^ T and NK cells. (**C**) Frequency of TCR clones projected on CD8^+^ T and NK cell UMAP and split by condition (SEV – severe, ASX – asymptomatic, CTRL – Control). (**D**) A boxplot illustrating the percent of expanded clones out of total expanded clones per cell subset in pregnant versus non-pregnant COVID-19 patients. (**E**) Gene and protein expression that characterize cell subset CD8T_3; hex bin plots with log(CPM). (**F**) TCR clones with specificity to SARS-CoV-2 projected onto CD8^+^ T and NK UMAPs. (**G**) Percent of TCR clones with specificity to SARS-CoV-2 epitopes out of total cells in each cell subset (x-axis) and number of cells with SARS-CoV-2 specificity (annotation at the end of the bars). (**H**) Hill diversity index for various q values, every line represents a patient and is colored by condition. NonPreg_COVID includes non-pregnant patients with asymptomatic and severe COVID, Preg_COVID includes pregnant patients with asymptomatic and severe COVID. *p < 0.05, **p < 0.01, ***p < 0.001, p-values are shown after multiple hypothesis correction, if any test becomes not significant after multiple hypothesis correction it is denoted with ‘(ns)’ next to the unadjusted p-value (e.g.,*(ns)” refers to a p< 0.05 that became ns after multiple hypothesis correction).

Paired T cell receptor (TCR) repertoire sequencing was also analyzed for each of these specimens; expanded TCR clones were defined to have a frequency of at least 1% per sample and exist in at least 5 single cells. Using these parameters, we identified a total of 341 expanded clones across 10,833 single CD8^+^ T cells. Overall, the highest percentage of expanded clones were identified in the cytotoxic CD8T_2, the *CD40LG* expressing CD8T_3, and NK/NKT_1 cell subsets (**Figures S10A-D**). Within the context of SARS-CoV-2 infection, the frequency of expanded clones in pregnancy was significantly higher in the CD8T_3 cell subset compared to non-pregnant individuals, whereas clonal expansion of CD8 T cells occurred predominantly in CD8T_2 cells in non-pregnant individuals (**Figures 3C, D** and **Figures S10E–G**). The pregnancy-associated clonal expansion emerging from CD8T_3 – a subset of CD8 T cells with a distinctive transcriptional footprint characterized by low expression of *CD8A* and *CD8B*, and relatively high expression of *CD4, CD40LG,* and *CD6* (**Figure 3A, S6G**) – also demonstrated cytotoxic markers shared by neighboring cell subsets (*GZMH, GNLY, FGFBP2, PRF1*) and concurrent expression of surface protein markers CD45RO, KLRG1, CD150 (SLAM), and CD57 (**Figure 3E**). While similar cytotoxic CD4-expressing CD8^+^ T cells have been reported to be present in viral infections, including SARS-CoV-2^76,77^, our data highlighting the clonal expansion within this activated cytotoxic memory T cell population is novel in the context of pregnancy, indicating unique T cell clone expansion in pregnancy.

Using the database VDJdb^78^, we queried the identity of putative cognate epitopes to the TCR clones identified in our dataset and identified epitopes for 91 different clones from 810 single cells (**Table S4**). Clones with specificity to SARS-CoV-2 epitopes were most abundant in the Tc17-like CD8T_9 cell subset and MAIT cells (**Figures 3F** and **G, Table S6**). While there were non-SARS-CoV-2-related epitopes identified that could putatively bind these TCR clones, no enrichment to a specific non-SARS-CoV-2 epitope was detected when the analysis was stratified to COVID-19 severity or specific CD8 T cell subsets, even while accounting for HLA alleles (see **Methods**, **Table S7**). TCR diversity of CD8^+^ T cell repertoires were not significantly different between pregnant COVID-19 patients compared to non-pregnant COVID-19 patients (**Figure S10G** by Mann-Whitney test on q=0,1,2,3,4). We did not detect expanded TCR clones in CD4^+^ T cells nor expanded B cell receptor (BCR) clones in our single-cell cohort (**Figures S7A, D–F, S8A, D–F**). Taken together, these data demonstrate differences in CD8^+^ T cell adaptive host immune responses, TCR clonality, diversity, and potential to bind viral epitopes in COVID-19 infection during pregnancy.

### Impaired ISG expression in pregnant women

Interferon-stimulated gene (ISG) expression is a key immune response for protective SARS-CoV-2 immunity^79,80^. However, uncontrolled ISG responses may contribute to COVID-19 severity^81,82^. We assessed ISG expression across PBMC lineages in non-pregnant and pregnant patients with COVID-19, leveraging our scRNA-seq analyses to investigate how pregnancy influences IFN responses. Interestingly, pregnant individuals with COVID-19 had lower ISG expression across all PBMC lineages analyzed compared to non-pregnant individuals with COVID-19 (**Figure 4**). We found decreased expression signature of 99 ISGs (see list in **Table S5**) in CD8^+^ T cell subsets CD8T_1, 2, 3, 4, 6, 8 and NK/NKT cell subsets in pregnant compared to non-pregnant individuals with COVID-19 (**Figures 4A** and **B**). Similar results were obtained across MNP, CD4^+^, and B cell subsets defined by scRNA-seq (**Figures 4C–H**). While there was no significant difference in differential abundance in the eight MNP populations observed across either COVID-19 severity or pregnancy perturbation states (**Figures S9C** and **D**), in all MNP cell subsets (except pDCs) – including four CD14 high and CD16 negative cell subsets (MNP_1, MNP_2, MNP_3 and MNP_4), two cell subsets with varying expression of CD16 (MNP_5, MNP_ 6), and cDCs – there was decreased expression of ISGs in pregnant compared to non-pregnant patients with COVID-19 (**Figure 4D**). Interestingly, similar results were also identified in CD4^+^ T cell subsets (**Figure 4F**). While there were no significant differences in cluster abundance across the eight CD4 T cell subsets defined by scRNA-seq analysis (**Figures S7A–C**), there was decreased expression of ISGs in all CD4 expressing populations (**Figures 4E** and **F**). While ISG expression was not significantly different across B cell subsets (**Figures S8A–C**), there was significantly decreased abundance of the IFITM1, IFI44L expressing B cell subcluster 3 (B_3) in pregnant compared to non-pregnant patients with COVID-19 (**Figures 4G** and **H**), consistent with the theme of diminished ISG-related responses in pregnant individuals across multiple PBMC lineages. We also noted that, unlike non-pregnant individuals, plasma IFN-α 2 level was not increased in pregnant women even after SARS-CoV-2 infection (**Figure S11**), further indicating that the IFN response may be subdued in pregnancy. Reduced ISG responses in pregnant individuals with SARS-CoV-2 infection suggests one potential mechanism underlying pregnant individuals’ increased susceptibility to severe viral infections.

**Figure 4.**
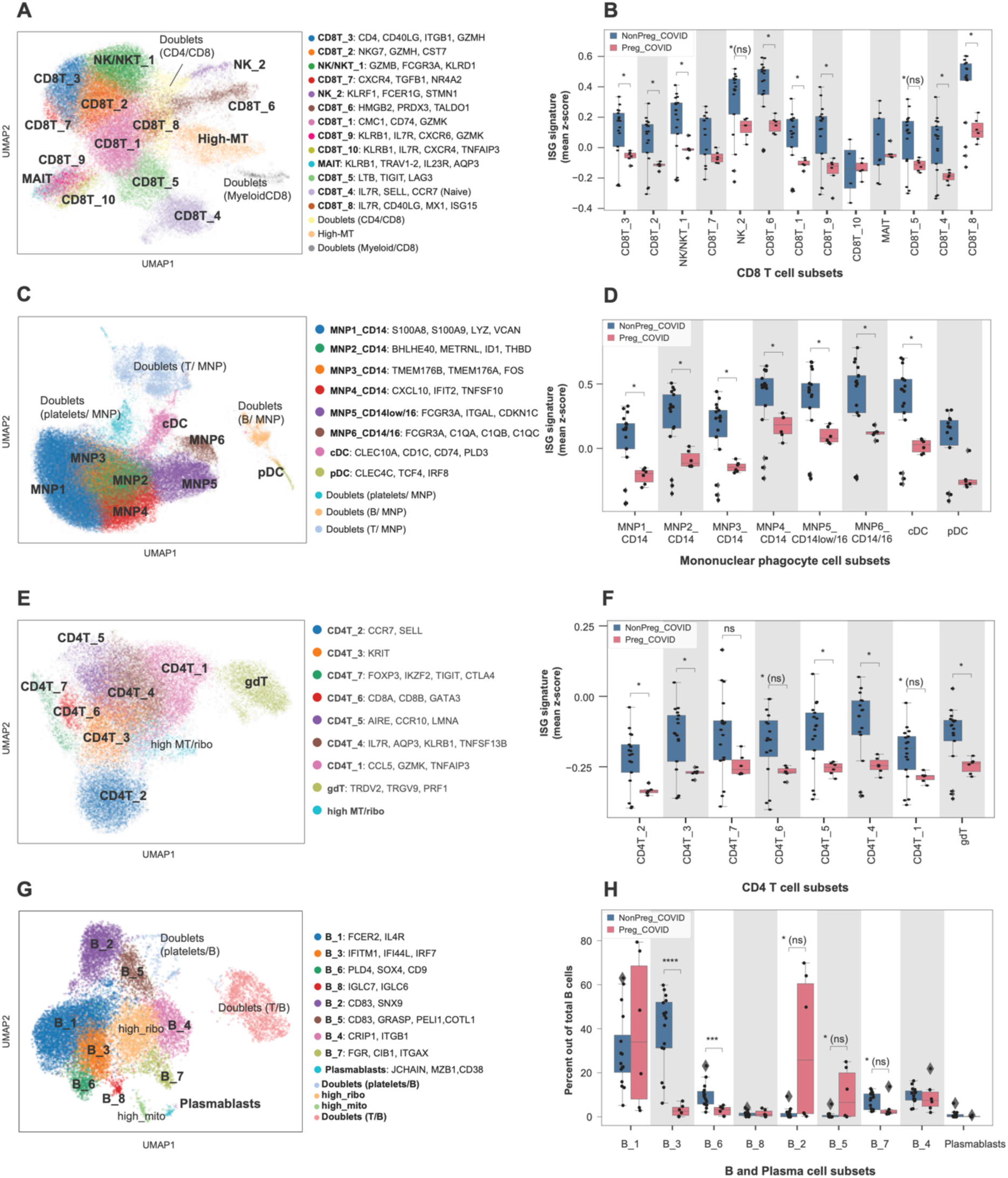
Interferon-stimulated genes are downregulated in pregnant patients with COVID-19. (**A**) Uniform Manifold Approximation (UMAP) embedding of 50,140 CD8^+^ T and NK cells and their clustering to 13 cell subsets. (**B**) A boxplot illustrating the expression signature of 99 interferon-stimulated genes (ISGs) across CD8^+^ T cell subsets in pregnant versus non-pregnant COVID-19 patients. (**C**) UMAP embedding of 66,719 mononuclear phagocyte (MNP) cells and their clustering to eight cell subsets. (**D**) A boxplot illustrating the expression signature of 99 ISGs across MNP cell subsets in pregnant versus non-pregnant COVID-19 patients. (**E**) UMAP embedding of 29,960 CD4^+^ T cells and their clustering to eight cell subsets. (**F**) A boxplot illustrating the expression signature of 99 ISGs across CD4 T cell subsets in pregnant versus non-pregnant COVID-19 patients. (**G**) UMAP embedding of 19,168 B and plasma cells and their clustering to nine cell subsets. (**H**) A boxplot illustrating the B cell subset abundances in pregnant versus non-pregnant COVID-19 patients. NonPreg_COVID includes non-pregnant patients with asymptomatic and severe COVID-19, Preg_COVID includes pregnant patients with asymptomatic and severe COVID-19. *p < 0.05, ***p < 0.001, p-values are shown after multiple hypothesis correction, if any test becomes not significant after multiple hypothesis correction it is denoted with ‘(ns)’ next to the un-adjusted p-value.

### Distinctive gut microbiota species in pregnant women with COVID-19

Growing evidence suggests that gut microbiota composition dictates the duration and magnitude of host immune responses upon respiratory infection^83^. Additionally, recent data indicate that SARS-CoV-2 infection induces changes in the gut microbiota^84^ and these changes may affect the disease course^85^. We performed shotgun metagenomic sequencing analyses with pregnant women’s stool samples [Pregnant controls (n = 29), Pregnant COVID-19 (n = 27) (13 mild, 6 moderate, 8 severe/critical), Non-pregnant COVID-19 (n = 13)] collected from SARS-CoV-2 negative and SARS-CoV-2-infected groups. We did not find differences in alpha diversity (a measure for species richness and evenness) (**Figure S12A**), and beta diversity (a statistic used to quantify the compositional dissimilarity) of microbial communities from SARS-CoV-2-infected pregnant women compared to those from non-pregnant women (**Figure S12B**). However, by investigating species-level differences, we found that pregnant patients with COVID-19 have an increased relative abundance of *Roseburia faecis*, *Eubacterium species CAG 38*, *Eubacterium eligens*, and a decreased abundance of *Lactobacillus rhamnosus* and *Erysipelatoclostridium ramosum* compared to patients with non-pregnant COVID-19 (**Figure S12C**). Among SARS-CoV-2-infected pregnant women with different clinical severity, we did not observe significant differences in alpha diversity according to control or case severity categories among pregnant women (**Figure S12D**) and beta diversity (**Figure S12E**). However, when we compared the bacterial species-level differences between pregnant patients with COVID-19 to pregnant controls, we found that *Phascolarctobacterium faecium*, *Parabacteroides merdae*, *Intestinimonas butyriciproducens*, *Bacteroides fragilis*, *Asaccharobacter celatus*, *Alistipes putredinis*, and *Adlercreutzia equolifaciens* were significantly reduced in pregnant patients with COVID-19 compared to pregnant controls. In parallel, *Actinomyces graevenitzii* increased in its abundance (**Figure S12F**). These data indicate that pregnancy influences the relative abundance of certain gut bacterial species, and alterations in gut microbiota are further affected by COVID-19 infection. Recent studies, including one from our group, indicated that the maternal microbiota community influences the long-lasting health of offspring^86–88^, thus, warranting future mechanistic studies of altered microbiota in pregnancy at baseline and upon viral infection.

### Distinct cytokine and chemokine responses in non-pregnant versus pregnant patients with COVID-19

The unrestrained production of cytokines or chemokines, the so-called “cytokine storm,” is a well-described feature in some cases of severe COVID-19^5,30,33,89^. To gain insight into the differences in inflammatory response between pregnant and non-pregnant individuals with COVID-19, we collected plasma samples from 107 individuals, including 38 pregnant patients with acute COVID-19, 31 non-pregnant with acute COVID-19, 21 SARS-CoV-2 negative uninfected pregnant controls, and 17 uninfected non-pregnant controls to assess the levels of 71 cytokines, chemokines, and growth factors (**Figure 5 and Figure S13**). We found that COVID-19 disease severity^47,48^ was a key consideration in understanding cytokine elevation or suppression in the setting of acute disease. Moreover, we identified substantial differences between the pregnant and non-pregnant cytokine/chemokine and growth factor response to COVID-19, with several different patterns of cytokine expression. Among those tested, only one cytokine, IL-10, was altered similarly in pregnant and non-pregnant individuals with acute COVID-19 infection; IL-10 increased with acute infection in both groups, consistent with earlier data from non-pregnant individuals^5,30,31,34^. In contrast, several cytokines, chemokines and growth factors were altered by COVID-19 in opposite directions between pregnant and non-pregnant individuals, including TNF-α, VEGF-A, CCL3, CCL4, CCL22, CCL5, and PDGF-AA/BB **(Figure 5)**; all of these were increased in acute COVID-19 in non-pregnant individuals but reduced in pregnant individuals with mild COVID-19, relative to respective uninfected control populations. A slight exception was TNF-α, which was increased in non-pregnant individuals with COVID-19 relative to uninfected controls and reduced among pregnant with mild COVID-19 but increased in pregnancy as disease severity increased. Many cytokines and chemokines were increased by pregnancy itself (**Figure 5)**, with significantly increased levels noted in pregnant uninfected controls relative to non-pregnant uninfected (IL-27, CCL3, CCL4, CCL22, CCL5, and PDGF-AA/BB). Finally, we noted several cytokines were significantly reduced in pregnant individuals with COVID-19 relative to non-pregnant individuals with COVID-19, including CCL2, CCL8, VEGF-A, CCL-3, CCL-4, CCL-22, CCL-5 and PDGF-AA/BB. In contrast, IL-27 was the only cytokine that was significantly increased among all pregnant groups (uninfected controls and COVID-19 acute infections) relative to their non-pregnant counterparts, and significantly suppressed in COVID-19 infection among pregnant patients, relative to pregnant controls^5,30,31,34,91,92^.

**Figure 5.**
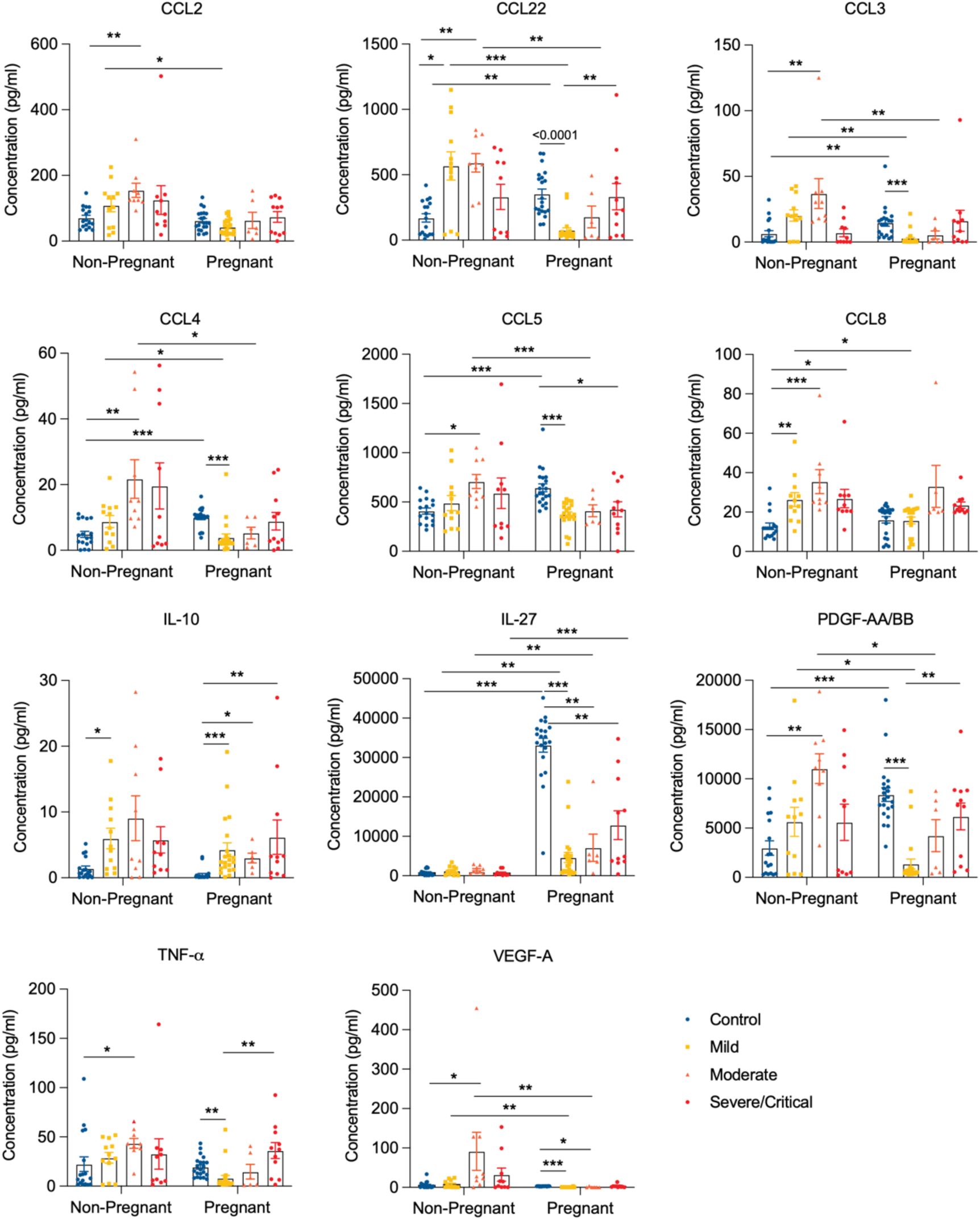
Comparative analyses of plasma cytokines, chemokines, and growth factors of pregnant versus non-pregnant control and COVID-19 patients. (**A**–**C**) Concentrations of cytokines, chemokines, and growth factors in pregnant and non-pregnant women who were healthy (Non-pregnant n = 17, Pregnant n = 21) or who had Mild (Non-pregnant n = 12, Pregnant n = 21), Moderate (Non-pregnant n = 9, Pregnant n = 6), or Severe/Critical (Non-pregnant n = 10, Pregnant n = 11) COVID-19. Data are shown as the mean ± SEM. *p < 0.05, **p < 0.01, ***p < 0.001.

Taken together, these results demonstrate the impact of pregnancy on baseline levels of immune-related soluble factors, such as cytokines and chemokines, in blood circulation. Their relative abundance in non-pregnant and pregnant individuals is further shaped by viral infection, indicating a complex interplay between the host immune response associated with pregnancy and respiratory viral infection that culminated in the altered production of cytokines and chemokines in blood circulation.

### High body mass index (BMI) and pregnancy synergistically alter immune responses

Obesity represents the most common comorbidity during pregnancy and has been recognized as a risk factor for severe COVID-19^90^. For example, obesity-associated, chronic, low-grade inflammation and impaired lung function may underlie enhanced susceptibility to infection^91^. Thus, we sought to examine whether maternal obesity (pre-pregnancy BMI ≥30 kg/m^2^) was associated with altered immune responses in pregnant patients with COVID-19, and whether obesity impacted immune responses to COVID-19 differently in pregnant versus non-pregnant populations. To do so, we grouped non-pregnant and pregnant patients with COVID-19 based on obesity status. Upon *in vitro* stimulation, CD4^+^ and CD8^+^ T cells produced higher levels of IL-2, IFN-γ, and TNF-α in pregnant individuals with obesity and COVID-19, compared to non-obese (BMI <30 kg/m^2^) pregnant individuals with COVID-19 (**Figures S14A** and **14B**). COVID-19- associated IL-4 production by CD4^+^ and CD8^+^ T cells was augmented by obesity, albeit not statistically significantly, in pregnant individuals. Thus, obesity had a more pronounced impact on enhanced T cell production of inflammatory cytokines in pregnant individuals with COVID-19 compared to non-pregnant individuals (**Figures S14A** and **14B**). Obesity augmented IFN-α plasma levels more significantly in pregnant women with COVID-19 compared to non-pregnant women, although IFN-α levels were significantly higher in non-pregnant individuals with COVID-19 compared to pregnant, regardless of obesity status (**Figure S14C**). Similar trends were observed for IL-1α and IL-17A, where obesity was associated with augmented plasma levels in pregnant women with COVID-19, although neither finding achieved statistical significance (*p*= 0.07 and *p*=0.05, respectively, **Figure S14C**). Obesity is known to be associated with suppressed monocyte function in pregnancy^92^, and with reduced monocyte numbers, monocyte exhaustion, and overall depressed innate immune function in the setting of COVID-19 infection in non-pregnant individuals^93^. In our analyses, however, obesity was not associated with further depression of the profound COVID-19-associated suppressed production of cytokines by monocytes. Taken together, these data suggest that obesity may increase the risk of COVID-19 in pregnant individuals by augmenting T cell cytotoxic and inflammatory functions, and that the profound suppression of monocyte function that occurs in COVID-19 may not be further augmented by obesity.

### Effect of fetal sex on immune responses in pregnant patients with COVID-19

While fetal sex was recently shown to influence COVID-19-related immune responses in the placenta^94^, it is unknown whether it also affects maternal immune responses. We first compared T cell composition and cytokine production based on fetal sex. Overall frequencies of naïve, CM and EM CD4^+^ and CD8^+^ T cells (**Figures S15A** and **S15B**) in mothers, and maternal cytokine production by CD4^+^ T cells (**Figure S15C**), CD8^+^ (**Figure S15D**), and γδT cells (**Figure S15E**), were comparable regardless of fetal sex. Mothers with severe or critical COVID-19 carrying a male fetus demonstrated reduced levels of plasma IL-10, IL-1RA and IL-27 relative to mothers carrying a female fetus, although none of these reductions achieved statistical significance (**Figure S15F**). Among pregnant individuals with moderate COVID-19 disease severity carrying a male fetus, we detected reduced production of IFN-λ IFN-β, and IL-6 by CD14^+^ monocytes upon *in vitro* LPS stimulation, relative to individuals with moderate COVID-19 carrying a female fetus (**Figure S15G**), although only the reduction in IFN-λ achieved statistical significance. Overall, these findings suggest that fetal sex may subtly influence maternal immune responses in the setting of SARS-CoV-2 in pregnancy, and this area is worthy of study in cohorts with larger numbers of males and females per biological group and/or a less heterogeneous viral infection phenotype.

### Dissecting the modulatory roles of IL-27 in maternal immune responses using respiratory viral infection and immune challenge models in mice

Plasma cytokine analyses revealed that IL-27 was highly expressed in healthy pregnant women and dramatically decreased in pregnant COVID-19 cases (**Figure 5**). In addition, a principal component analysis (PCA) for plasma cytokines identified IL-27 as a major driver in separating healthy pregnant women from other groups (**Figure S16A**). IL-27, a heterodimeric cytokine composed of p28 and Epstein-Barr virus-induced gene 3 (EBI3), has been implicated in both promoting and mitigating immune responses by augmenting Th1 and Treg cell responses, respectively^95,96^. IL-27 is also known to suppress macrophage and dendritic cell function and reduce the production of TNF-α^97,98^. While IL-27 levels in blood circulation have been previously linked to pregnancy-associated preeclampsia and preterm birth^99–101^, its role in pregnancy is not well understood.

We employed preclinical mouse models of influenza viral infection and polyinosinic: polycytidylic acid (poly[I:C))-dependent fetal demise to characterize IL-27 function in mice *in vivo*. Neither syngeneic nor allogeneic pregnant mice were found to have increased IL-27 levels at baseline (**Figure 6A**). However, consistent with an earlier report^102^, anti-CD3 injection led to robust induction of IL-27 in mice (**Figure S16B**). These data indicate that IL-27 is robustly induced in circulation in humans but not in mice during pregnancy. Therefore, we used a gain-of-function approach to determine the impact of high levels of IL-27 on viral infection severity and pregnancy outcome. We infected non-pregnant mice with a sub-lethal dose of influenza virus Puerto Rico 8 (PR8) strain, followed by supplementing them with daily injections of PBS or IL-27 (**Figure 6B**). Compared to the PBS-treated control, the IL-27-injected infected group suffered from significant weight loss and reduced survival (**Figures 6C** and **7D**). Additionally, we found a significantly elevated expression of lung damage-associated genes, such as *Muc5ac*, *F3*, and *Retnla* (**Figure 6E**), accompanied by increased albumin levels (another marker for lung damage) in bronchial-alveolar lavage fluid (BALF) of the IL-27-treated group (**Figure 6F**). Consistent with these findings, we detected enhanced viral gene (*Ns1*, *Np*) expression in the IL-27-treated mouse lung tissues (**Figure 6G**). Next, we induced inflammation in pregnant mice by intraperitoneally injecting them with high amounts of poly(I:C), an agent known to induce fetal demise^103,104^. We then treated them with PBS or IL-27 before monitoring the fetal survival rate at embryonic day 17.5 (E17.5) (**Figure 6H**). Compared to PBS control, IL-27 supplementation significantly enhanced fetal survival without altering total fetal numbers (**Figures 6I** and **J**).

**Figure 6.**
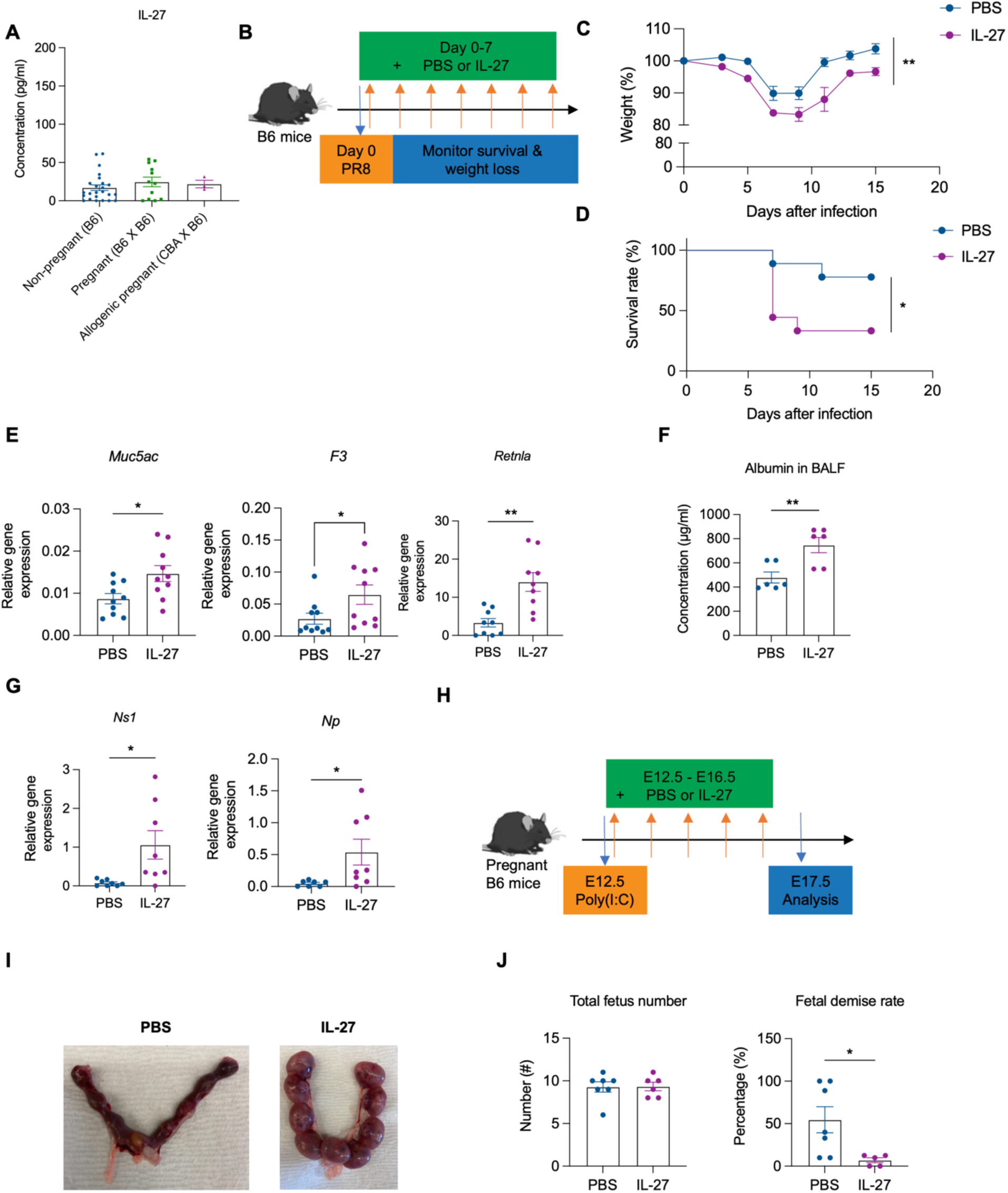
IL-27 protects developing fetuses from maternal inflammation. (**A**) IL-27 protein levels in the plasma of non-pregnant, E17.5 syngeneic (B6 x B6) and allogenic (CBA x B6) mice measured by ELISA. (**B**) Experimental scheme of IL-27 gain-of-function model. On day 0, mice were infected with 50 CFUs of PR8 intranasally, followed by intraperitoneal administration of PBS or 500 ng of recombinant IL-27 from day 0 to day 7. (**C** and **D**) Weight loss and survival rate were monitored in PBS (n = 9) and IL-27-treated mice (n = 8). (**E**) Three days post PR8 infection, the expressions of lung damage-associated genes were determined using RT-qPCR. (**F**) In three days post PR8 infection, the concentration of albumin in BALF was measured with albumin ELISA. (**G**) Influenza viral genes in mouse lungs were measured with RT-qPCR. Gene expression was normalized with beta-actin. (**H**) Experimental scheme of IL-27 gain-of-function experiment of poly (I:C)-induced fetal demise model. On day E12.5, mice were infected with poly(I:C) and then intraperitoneally injected with PBS or 1000 ng of recombinant IL-27 from E12.5 to day 16.5. (**I** and **J**) Representative images of fetuses from PBS or IL-27-treated pregnant dams (**I**) on E17.5. Total number of fetuses and fetal demise rate (non-viable/resorbed fetus numbers/total fetus numbers X 100) were measured in PBS or IL-27-treated pregnant mice (**J**). Data are shown as the mean ± SEM. *p < 0.05, **p < 0.01.

These convergent results, from two different mouse models, indicate that enhanced levels of IL-27 help protect developing fetuses from uncontrolled inflammation while making mice more vulnerable to viral infection. Thus, elevated IL-27 levels during pregnancy in humans may act as a double-edged sword: beneficial in protecting developing fetuses from SARS-CoV-2-induced inflammation while potentially exposing pregnant women to viral inflammation-associated pathology.

## Discussion

Studies have suggested that pregnant women are at a higher risk of experiencing complications and mortality associated with SARS-CoV-2 infection^105–107^. However, the mechanisms underlying this increased susceptibility during pregnancy are still not well understood. Furthermore, conflicting findings in earlier studies, perhaps due to limitations such as small sample sizes, heterogeneous timing of sample collection, and/or lack of availability of appropriate pregnant and non-pregnant controls, posed a challenge in formulating a comprehensive understanding of the modulatory effects of pregnancy on immune responses to SARS-CoV-2 and other similar viral infections.

To gain deeper insights into how pregnancy affects host immune responses at baseline, and in the presence of viral infections, we conducted a study profiling a wide range of immune sample types including PBMC, peripheral plasma/sera, and the gut microbiome (stool), and employing diverse immunological strategies. We characterized samples collected from pregnant and age-matched non-pregnant women during the acute and convalescent phases of SARS-CoV-2 infection, compared to samples from pregnant and non-pregnant SARS-CoV-2-negative controls.

Earlier work suggested that the activation of CD4^+^ and CD8^+^ T cells appears to be similar between pregnant and non-pregnant women^41^. In COVID-19 pregnant women, we observed impaired CD8^+^ T cell responses characterized by lower percentages of EM cells, reduced cytokine expression, reduced ISG expression, and altered clonal expansion compared to COVID-19 non-pregnant women. CD4-expressing CD8^+^ T cells in pregnant patients with COVID-19 were found to undergo a clonal expansion, while there was a clonal expansion of another cytotoxic CD8+ T cells in non-pregnant patients with COVID-19. Additionally, CD8^+^ T cells in infected pregnant women displayed reduced functionality and signs of T cell exhaustion. We also identified decreased anti-inflammatory Treg percentages in pregnant patients with severe/critical COVID-19 compared to non-pregnant patients. These data suggest that pregnancy-associated CD8^+^ T cells may be less equipped to combat viral infections effectively due to their higher expression of PD-1 and Tim-3, lower EM cell percentages, altered clonal expansion, and reduced Treg levels.

Impaired cytokine production by monocytes has been reported in severe cases of COVID-19^108,109^. However, the specific functional changes in pregnant monocytes compared to non-pregnant monocytes during COVID-19 were unknown. We found that pregnancy is associated with enhanced monocyte function at baseline compared to non-pregnant women. Monocytes from pregnant women exhibited increased HLA-DR expression, cytokine production, and chemokine production. This enhanced monocyte activity might compensate for impaired T cell function associated with pregnancy, as monocytes play a crucial role in recruiting leukocytes to inflammatory sites through the production of chemokines, such as CCL3, CCL4, and CCL22^110^. Our data are consistent with other reports that monocyte function in pregnancy may be enhanced relative to the non-pregnant state^111,112^. However, we also noted that SARS-CoV-2 infection led to long-lasting suppressive effects on monocytes that were most pronounced in pregnant women, demonstrated by reduced HLA-DR expression in intermediate and non-classical monocytes persisting into convalescence, dampened cytokine production in severe/critical disease, and reduced ISG expression in pregnant compared to non-pregnant individuals with COVID-19. In addition, given the heightened monocyte function we observed in uninfected pregnant individuals, the COVID-19-associated monocyte suppression marks an even more profound difference from the pregnant baseline state than does monocyte suppression observed in non-pregnant individuals with COVID-19^109,113^.

Our data indicated substantial differences between the pregnant and non-pregnant cytokine/chemokine and growth factor response to COVID-19. We also noted significantly reduced IFN-α levels in pregnant individuals (both with and without COVID-19). Consistent with these findings, PBMCs from pregnant patients with COVID-19 were found to have a dampened ISG signature compared to non-pregnant counterparts, providing another possible mechanism for enhanced susceptibility to severe viral infection. We also found that pregnant patients with COVID-19 carry different bacterial species compared with pregnant uninfected individuals, as well as non-pregnant patients with COVID-19. Recent studies, including one from our group, indicated that the maternal microbiota community influences the long-lasting health of offspring^86–88^, warranting future mechanistic studies of altered microbiota in pregnancy. Finally, we also demonstrated that CD4^+^ and CD8^+^ T cells from COVID-19–infected pregnant individuals with obesity produced increased cytokine levels upon stimulation compared to non-obese pregnant individuals with COVID-19. SARS-CoV-2 infection led to aggravated inflammatory cytokine responses only in pregnant subjects with obesity. Such interaction between obesity and COVID-19 status was not observed in non-pregnant groups. The enhanced inflammatory response may contribute to a higher risk of developing inflammatory pathologies in pregnant women with obesity. Using unbiased cytokine analyses and preclinical mouse models, we identified IL-27 as a pregnancy-associated cytokine and functionally determined its role in influencing maternal immune responses and fetal survival. To build upon the finding of significantly elevated levels of IL-27 in pregnant individuals with and without COVID-19, we used mouse models to demonstrate that administering IL-27 to mice with respiratory viral infection resulted in more severe lung damage and increased mortality while simultaneously being associated with protection of the fetuses under poly(I:C)-induced inflammation during pregnancy. This suggests the potential for a tradeoff between fetal survival and severe maternal morbidity in the setting of specific cytokine responses to viral infection. IL-27 was previously shown to drive PD-L1 and Tim-3 expression and underlie T cell dysfunction^45,114^. Given that we found PD-1 and Tim-3 expression to be increased in pregnancy and in pregnant women with COVID-19, an important direction for future research will be to dissect the role that enhanced IL-27 expression in pregnancy might play in pregnancy-specific T cell exhaustion phenotypes. In addition, IL-27 plays an important role in protection from exaggerated inflammation, but high levels of IL-27 at early time points during infection impaired virus clearance and worsened disease in a murine influenza virus infection model^115^. Further studies are needed to investigate the pleiotropic role and pregnancy-specific role of IL-27.

There are limitations to our study. First, our COVID-19 pregnant samples were largely from third trimester infections because these samples were collected either upon hospital admission for symptomatic COVID-19 infection (more likely for patients to have severe infection requiring hospitalization in the third trimester of pregnancy compared to first or second), or upon presentation to labor and delivery for labor or other pregnancy-related evaluation (universal SARS-CoV-2 status screening was conducted with SARS-CoV-2 nasopharyngeal PCR from April 2020 onward). Given hospital restrictions on research-only study visits at our hospitals and most across the country, it was not possible to comprehensively sample first and second trimester pregnant patients for SARS-CoV-2 infection, so neither our nor any specimens banked contain a comprehensive sampling of first and second trimester SARS-CoV-2 infection. Given that trimester-specific immune signatures have been described^116,117^, how SARS-CoV-2 infection in the first and second trimesters affects maternal host immune responses is a question that remains to be answered, but may not be answerable with existing datasets. Second, we evaluated non-specific T cell responses during the acute and convalescent phases of COVID-19 without assessing SARS-CoV-2-specific T cell responses. Another important area for future study beyond the scope of the experiments performed here is to understand how maternal immune responses during acute SARS-CoV-2 infection and convalescence relate to longer-term immune function in individuals after pregnancy^118,120,122^ (e.g., development of long COVID or other immune syndromes), and the short- and long-term health outcomes of their offspring. Members of our study team have examined both short-term neurodevelopmental^119,121^ and cardiometabolic^123,124^ outcomes in offspring of mothers infected with SARS-CoV-2 in pregnancy, demonstrating increased risk for neurodevelopmental diagnoses at 12 months particularly pronounced in male offspring, and increased risk for altered growth trajectories in the first year of life that may presage an increased risk for early-onset cardiometabolic complications. These follow-up studies did not link maternal immune responses 1:1 with offspring health/immune trajectories, however, and establishment of prospective cohorts to link maternal immune activation with the neurodevelopmental and cardiometabolic risk of individual offspring remains an important area for future study^25^.

In summary, we report the the largest and most comprehensive immune survey of COVID-19 in pregnancy to date, and we identified pregnancy-specific alterations in immune responses to SARS-CoV-2, including altered T cell and monocyte function. Our findings of increased T cell exhaustion and profound monocyte suppression with infection in pregnancy relative to uninfected pregnant individuals, a pregnancy-specific unique clonal expansion of CD4-expressing CD8^+^ T cell subsets, and reduced expression of interferon-stimulated genes by all pregnant immune cell subsets suggest potential mechanisms by which pregnancy-specific immunity predisposes individuals to increased severity of disease with viral infection. Furthermore, through probing our insights from human cytokine data in a murine model, we identified IL-27 as a key cytokine that underlies pregnancy-associated immune changes that predispose the mother to severe respiratory disease while protecting the fetus from demise. Altogether, this work elucidates novel key aspects of pregnancy immunity that are important not only in understanding pregnancy-specific responses to SARS-CoV-2 but with broad implications for a novel understanding of altered immunity and enhanced susceptibility to viral infection in pregnancy that can be used to guide targeted screening and therapeutic strategies in future pandemics.

## Data Availability

scRNA data, CITE-seq, and repertoire data were deposited to GEO and will be available in the accession number GSE239452 upon publication. The code used for single-cell analysis and figures generation will be available in GitHub at https://github.com/villani-lab/COVID_pregnancy_and in the Zenodo repository upon publication of this study.

## STAR Methods

### EXPERIMENTAL MODEL AND SUBJECT DETAILS

#### Human participants and data collection

Pregnant women receiving care at Massachusetts General Hospital and Brigham and Women’s Hospital in Boston, MA, and enrolled in a COVID-19 pregnancy biorepository between April 2020 and January 2021, prior to the widespread availability of COVID-19 vaccines, were included in this study (MGB IRB#2020P003538 and #2020P000804). Pregnant women were eligible for inclusion if they were 18 years of age or older, able to provide informed consent for themselves or with a healthcare proxy, and diagnosed with, or at risk for SARS-CoV-2 virus infection. All participants were tested for the presence of SARS-CoV-2 by RT-PCR of nasopharyngeal swab on admission to Labor and Delivery and/or for symptoms of COVID-19. Contemporaneous uninfected pregnant individuals (again status confirmed by SARS-CoV-2 RT-PCR of nasopharyngeal swab) were also enrolled at delivery. Individuals with autoimmune disease/use of immune-modulating medications, and those with other infections, such as clinical chorioamnionitis, or other symptoms of viral or bacterial infection were also excluded.

Non-pregnant women hospitalized with confirmed SARS-CoV-2 infection and enrolled contemporaneously as part of a general adult cohort described previously^125^ (MGB IRB #2020P000804) were used as an additional comparator group. Banked PBMC and plasma samples from healthy, non-pregnant, control women collected prior to the COVID-19 pandemic were used as an additional biological comparator group. These participants were known to be negative for HIV, TB, EBV, Hepatitis B and C, and Dengue virus, with normal complete blood counts and not on any immune-modulating medications; their samples were collected as part of a protocol (MGB IRB # 2010P002121) to bank healthy control samples for use as biological comparators in infectious disease studies.

#### Mice

All animals were housed in individually ventilated cage systems (Tecniplast) in a specific pathogen-free facility maintained at 20–22 °C with 40–55% humidity and under a 12-h light/12-h dark cycle. All experiments were conducted in accordance with procedures approved by the Institutional Animal Care and Use Committee of Harvard University.

### METHOD DETAILS

#### Clinical and sample definitions

Clinical and demographic information were abstracted from the electronic medical record. COVID-19 symptom severity was determined based on NIH criteria^47,48^. Samples not designated “convalescent” were collected from pregnant participants in closest proximity to acute COVID-19 illness and within 30 days of a positive SARS-CoV-2 test. Samples from uninfected pregnant participants, and from convalescent individuals who tested positive for SARS-CoV-2 infection during pregnancy but remote (>30 days) from delivery sample collection, were collected during the delivery hospitalization on admission to Labor and Delivery. Details on sample collection procedures have been described previously^126^.

#### Blood specimen processing

Blood samples were obtained by venipuncture into EDTA tubes. Blood was centrifuged at 1000 × *g* for 10 mins, and plasma was aliquoted and stored at -80 °C. PBMCs were collected using a Ficoll density gradient^127^. Briefly, blood was transferred to a 50 mL conical tube and diluted 1:1 with Hanks’ balanced salt solution (HBSS) without calcium or magnesium. Diluted blood was layered on top of Ficoll in a 2:1 ratio. The conical tube was then centrifuged at 1000 × *g* for 30 min at room temperature without braking. The interphase was collected and placed in a new 15 mL conical tube with HBSS to bring the volume to 15 mL and centrifuged at 330 × *g* for 10 mins with high brake. The supernatant was removed and the cell pellet washed with HBSS. Cells were frozen in freezing medium with RPMI 1640 medium with 1% penicillin-streptomycin, L-glutamine, 1% sodium pyruvate, 1% non-essential amino acids, 20% FBS, and 10% DMSO at 5–10 million cells/cryovial, placed in a chilled Mr. Frosty, stored at -80 °C for one day, then transferred to liquid nitrogen for long-term storage.

#### Flow cytometry-based immune cell profiling

PBMCs were plated at 1 × 10^6^ cells per well in a 96-well U-bottom plate. PBMCs were stimulated with eBioscience™ Cell Stimulation Cocktail (plus protein transport inhibitors) (Thermo Fisher Scientific) in RPMI (RPMI-1640 supplemented with 10% fetal bovine serum (FBS), 2 mM L-glutamine, 100 U/ml penicillin, and 100 mg/ml streptomycin, 1 mM sodium pyruvate, and 50 μM 2-mercaptoethanol) for 5 hrs to monitor cytokine expression. After stimulation, cells were stained with antibodies against cell-surface markers and the LIVE/DEAD fixable dye Aqua (Thermo Fisher Scientific) to exclude dead cells, fixed and permeabilized with a FoxP3 transcription factor staining kit (Thermo Fisher Scientific), and subsequently stained with cytokine- and/or transcription factor-specific antibodies. All flow cytometry analyses were performed on a Symphony flow cytometer (BD), and data were analyzed with FlowJo software (BD). The specific markers used to identify each subset of cells are summarized in the gating strategy (**Figure S1**). Antibodies that were used for flow cytometry are listed in the Table below.

| **Table Antibodies** | **Source** | **Identifiers** |
| --- | --- | --- |
| Anti-Human CD3 AF700 | Thermo Fisher Scientific | Cat# 56-0038-82 |
| Anti-Human CD4 PE-Cy5 (Clone RPA-T4) | BioLegend | Cat# 300510 |
| Anti-Human CD8 PE-Cy5.5 (Clone RPA-T8) | Thermo Fisher Scientific | Cat# 35-0088-42 |
| Anti-human CD19 BUV496 (Clone SJ25C1) | BD | Cat# 612938 |
| Anti-human HLA-DR FITC (Clone Tü36) | BioLegend | Cat# 361604 |
| Anti-human PD-1 BUV615 (Clone EH12.1) | BD | Cat# 612991 |
| Anti-human FOXP3 PE (Clone 206D) | BioLegend | Cat# 20108 |
| Anti-human TIM-3 BUV737 (Clone 7D3) | BD | Cat# 748820 |
| Anti-human CD25 BV650 (Clone BC96) | BioLegend | Cat# 302634 |
| Anti-human TCR-$\gamma\delta$ BUV563 (Clone 11F2) | BD | Cat# 748534 |
| Anti-human IFN-γ BV750 (Clone B27) | BD | Cat# 566357 |
| Anti-human TNF-α FITC (Clone Mab11) | BioLegend | Cat# 502906 |
| Anti-human IL-2 BV605 (Clone JES6-5H4) | BioLegend | Cat# 503829 |
| Anti-human IL-4 BV786 (Clone MP4-25D2) | BD | Cat# 564113 |
| Anti-human Granzyme B BV421 (Clone QA18A28) | BioLegend | Cat# 396413 |
| Anti-human IL-17A PE-Cy7 (Clone eBio64DEC17) | Thermo Fisher Scientific | Cat# 25-7179-42 |
| Anti-human CD56 BV421 (Clone NCAM16.2) | BD | Cat# 562751 |
| Anti-human CD123 BUV805 (Clone 9F5) | BD | Cat# 742031 |
| Anti-human CD14 PE-Cy7 (Clone M5E2) | BioLegend | Cat# 301814 |
| Anti-human CD16 BUV786 (Clone 3G8) | BioLegend | Cat# 302046 |
| Anti-human CD16 BUV805 (Clone 3G8) | BD | Cat# 748850 |
| Anti-human CD45RO BUV661 (Clone UCHL1) | BD | Cat# 749887 |
| Anti-human CCR7 CF-594 (Clone 2-L1-A) | BD | Cat# 566768 |
| Anti-human CD38 BV750 (Clone HIT2) | BD | Cat# 746909 |
| Anti-human CD66b PerCP-Cy5.5 (Clone G10F5) | BioLegend | Cat# 305108 |
| Human BD Fc Block™ (Clone Fc1) | BD | Cat# 564219 |

#### Monocyte enrichment and stimulation

PBMC numbers were counted, and 2 x 10^6^ of PBMCs were used to enrich for CD14^+^ monocytes with MACS microbeads system (Miltenyi Biotec) according to the manufacturer’s protocol. 1 x 10^5^ cells of MACS-sorted CD14^+^ monocytes were stimulated with 1 ng/mL LPS in 200 μL of 10%FBS RPMI-1640 media for 8 hrs. After stimulation, cells were centrifuged, and supernatants were kept at -80 °C for the downstream analysis.

#### Cytometric bead array

Cytokine and chemokine concentrations were measured from the supernatants of CD14^+^ monocytes by cytometric bead array according to the manufacturer’s protocol (BioLegend).

#### Plasma cytokine and chemokine analysis

Sera were shipped to Eve Technologies (Calgary, Alberta, Canada) on dry ice, and levels of cytokine, chemokine, and growth factor were measured with Human Cytokine/Chemokine 71-Plex Discovery Assay (HD71). Serum levels of IL-1RA were validated by an IL-1RA Human ELISA Kit (Invitrogen).

#### Single-cell cohort details

Samples from pregnant participants collected as part of a healthcare-system-wide COVID pregnancy biorepository described previously^126,128^ were selected for single-cell sequencing to span a range of disease severity as follows: 3 samples from healthy uninfected pregnant “control” participants at term (39-40 weeks), 3 samples from asymptomatic pregnant SARS-CoV-2 positive individuals who tested positive at universal screening upon hospital admission (34, 39 and 40 weeks), and 3 samples from pregnant individuals PCR positive for SARS-CoV-2 with severe or critical COVID-19 as defined by NIH criteria^48^ (2 critically-ill at 34 and 41 weeks, 1 severe at 36 weeks). All included samples were collected prior to the initiation of any treatment.

The non-pregnant participants were selected from an MGH COVID-19 patient cohort described previously^129^. Out of this large cohort we chose the “non-pregnant severe” sample group by the following criteria: samples of female patients, 20–34 years old, PCR positive for SARS-CoV-2 with severe or critical COVID-19 by NIH criteria^48^, The “non-pregnant asymptomatic” sample group was selected by the following criteria: samples of female patients, 20–34 years old, PCR positive for SARS-CoV-2, without symptoms (none of these patients were admitted to the hospital within a 28 day window, all survived). All included samples were from day 0 before any treatment started. Further meta-data is available in Table S3.

#### PBMC CD45^+^ enrichment, cell hashing, CITE-seq staining

Cryopreserved PBMC samples (total of nine samples from the MGH pregnancy cohort and 17 samples from the MGH COVID-19 acute cohort) were thawed at 37 °C, diluted with a 10x volume of RPMI with 10% heat-inactivated human AB serum (Sigma Aldrich), and centrifuged at 300 × *g* for 7 mins. Cells were resuspended in CITE-seq buffer (RPMI with 2.5% [v/v] human AB serum and 2 mM EDTA) and added to 96-well plates. Dead cells were removed with an Annexin-V-conjugated bead kit (Stemcell Technologies, 17899) and red blood cells were removed with a glycophorin A-based antibody kit (Stemcell Technologies, 01738); modifications were made to the manufacturer’s protocols for each to accommodate a sample volume of 150 μL. Cells were quantified with an automated cell counter (Bio-Rad, TC20), after which, 2.5 x 10^5^ cells were resuspended in CITE-seq buffer containing TruStain FcX blocker (BioLegend, 422302) and MojoSort CD45 Nanobeads (BioLegend, 480030). Hashtag antibodies (BioLegend) were added to samples (**Table S9**) followed by a 30 mins incubation on ice. Cells were then washed three times with CITE-seq buffer using a magnet to retain CD45^+^ cells. Live cells were counted with trypan blue, and samples from each cohort bearing different hashtag antibodies were pooled together at equal concentrations.

For the pregnancy cohort, three pools of three samples across COVID-19 severity (severe COVID-19, asymptomatic SARS-CoV-2 infection, and no-SARS-CoV2 infection) were pooled (three pools) and loaded on three channels (total of nine channels), and the data from each donor and each channel was concatenated for computational analysis. For the MGH acute cohort, samples were pooled in groups of eight and loaded on two channels; individual donors selected as controls from the acute cohort were age- and gender-matched to the participants in the pregnancy cohort. Pooled samples were filtered with 40 μM strainers, centrifuged, and resuspended in CITE-seq buffer with TotalSeq-C antibody cocktail (BioLegend; **Table S9**). Cells were incubated on ice for 30 mins, followed by three washes with CITE-seq buffer and a final wash in the same buffer without EDTA (RPMI with 2.5% [v/v] human AB serum). Cells were resuspended in this buffer without EDTA, filtered a second time, and counted.

#### Single-cell gene expression, feature bar code, and TCR/BCR library construction and sequencing

An input of 50,000 hashed PBMC samples was loaded on a single channel in the 10X Chromium instrument for blood immune cell analysis, aiming for a recovery goal of 25,000 single cells. Hashed PBMC single-cell libraries were generated with the Chromium Single Cell 5’ kit (V2 NextGEM 10X Genomics, PN-1000263) together with the 5’ Feature Barcode library kit (10X Genomics PN-1000256). PCR-amplified cDNA was used for TCR and BCR enrichment with the Chromium Single Cell V(D)J Enrichment kit (10 Genomics PN-1000252 and PN-1000253). The library quality was assessed with an Agilent 2100 Bioanalyzer. All gene expression, feature barcode, TCR and BCR libraries were sequenced on an Illumina NovaSeq using the S4 300 cycles flow with the following sequencing parameters: read 1 = 26; read 2 = 90; index 1 = 10; index 2 =10.

#### scRNA-seq read alignment and quantification

##### RNA data

Raw sequencing data was pre-processed with CellRanger (v6.0.1, 10x Genomics) to demultiplex FASTQ reads, align reads to the human reference genome (GRCh38), and count unique molecular identifiers (UMI) to produce a cell x gene count matrix^130^. We used the Terra platform (https://app.terra.bio/) to run Cell Ranger via the workflow script that is part of a collection called Cumulus^131^. All samples were genetically demultiplexed using Souporcell^132^ (v2.0). Hashtag oligos (HTOs) were demultiplexed by Pegasus (v1.7.1, python) functions estimate_background_probs and demultiplex. Souporcells’ assignments were mapped to the demultiplexed HTOs of patients to identify the Souporcell assignment of the genetically demultiplexed samples. We used the results from the genetic demultiplexing because they resulted in fewer “doublet” and “unknown” assignments.

All count matrices were then aggregated with Pegasus using the aggregate_matrices function. Low-quality droplets were filtered out of the matrix prior to proceeding with downstream analyses using the percent of mitochondrial UMIs and number of unique genes detected as filters (<20% mitochondrial content, >300 unique genes). The percent of mitochondrial UMI was computed using 13 mitochondrial genes (MT-ND6, MT-CO2, MT-CYB, MT-ND2, MT-ND5, MT-CO1, MT-ND3, MT-ND4, MT-ND1, MT-ATP6, MT-CO3, MT-ND4L, MT-ATP8) using the qc_metrics function in Pegasus. The counts for each remaining cell in the matrix were then log-normalized by computing the log1p (counts per 100,000), which we refer to in the text and figures as log(CPM).

##### Protein data

Antibody-derived tags (ADT) data was demultiplexed by matching cell barcodes of the protein data to that of the RNA data cell barcodes. Only cells that had RNA data were paired with the protein data, we did not include cells that had only protein data without a matching RNA data. Protein data demultiplexing was therefore projected from the RNA-based demultiplexing. All protein values were CLR^133^ (centered log ratio) transformed by the following formula where x denotes an ADT vector of a single cell *c* and *G_c_* is the gene expression of all genes for cell *c*:

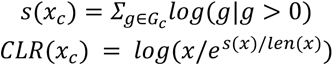

#### Basic and lineage-level single-cell clustering

All cells from all samples after the above-mentioned filtering were used for clustering. We identified candidates for highly variable genes as those that are expressed in at least 0.05% of the cells. Then, 2,000 highly variable genes (HVGs) were selected using the highly_variable_features function in Pegasus and used as input for principal component analysis. TCR and BCR genes were pre-excluded from the candidate HVGs. To account for batch effects, the resulting principal component scores were aligned using the Harmony algorithm^134^. The resulting principal components were used as input for Leiden clustering and Uniform Manifold Approximation and Projection (UMAP) algorithm. We identified main lineages using marker genes: T/NK cells (CD3E, CD3D, CD8A, CD8B, CD4, NCAM1, FCGR3A), B and plasma cells (MS4A1, CD19, JCHAIN) and MNPs (VCAN, LYZ, CD136, MARCO, CD14, HLA-DRB1, HLA-DRA). To further split CD4 T cells from CD8 T cells, we had to further split the T/NK subset by CD4, CD8, CD3E, CD3D, and NCAM1 (**Figures S6H** and **I**). In the resulting embedding and clustering (**Figure S6H**) clusters 4, 5, 7, 13, 16, and 17 were accounted as CD4 T cells and were re-sub-clustered (**Figure 4E**); clusters 1, 2, 3, 9, 10, 11, 12, 14, 15, 18 were accounted as CD8^+^ T cells (**Figure S6H**) and were re-sub-clustered (**Figure 3A**); and clusters 6 and 8 were accounted as NK cells (**Figure S6H**). Of note, some NK cells were found in the CD8 T cell lineage and were annotated as such. For every lineage separately we re-ran the clustering procedure: identified 1,000 HVGs, ran PCA, computed a UMAP embedding based on 50 PCs, and clustered using the Leiden algorithm.

#### Single-cell subset annotation

To annotate cell subsets at the lineage-level clustering, we used two complementary methods to identify marker genes and proteins: “one versus all” (OVA) in which we test which genes are differentially expressed between each cluster and the rest of the cells that do not belong to it; and “all versus all” (AVA) in which we test which genes are differentially expressed between every pair of clusters. For OVA, we calculated the area under the receiver operating characteristic (AUROC) curve for the log(CPM) values of each gene as a predictor of cluster membership using the de_analysis function in Pegasus. Genes with an AUROC ≥0.75 were considered as marker genes for a particular cell subset. In addition, we computed a pseudo-bulk count matrix by counting UMIs for every patient in every cluster. We then tested differential analysis using the Limma package (v3.54.2, R)^135^ and modeling gene ∼ is_clust using the lmFit() function, where is_clust is a factor with levels indicating if the sample was or was not in the subset being tested. For AVA, we used the pseudo-bulk matrix to model the gene expression in each cluster separately gene ∼ clust using the lmFit() function, where clust denotes the cluster, we then contrasted every cluster with every other cluster in this model using Limma’s contrasts.fit() function. The pseudo-bulk and AUROC statistics of OVA differentially expressed genes (DEGs) for all cell subsets can be found in **Table S8**.

Marker genes for each cell subset were interrogated and investigated for several different cluster resolutions using Cell Guide^136^. For each lineage, we identified the final resolution in which every cluster has a unique set of marker genes differentiating it from the rest of the clusters. If two cell clusters were indistinguishable, they were merged to create one cell subset to obtain the best possible resolution. The numbering of cell subsets in every lineage was finalized to be sorted by the cell subset size. The resolutions used for the final lineage clustering were as follows: T/NK cells resolution=1.8, CD4 T cells resolution=1, MNPs resolution=1, B/ Plasma cells resolution=1.3.

#### Single-cell differential abundance analysis

To test differential abundances in different cell subsets across conditions, we computed the abundance of each cell subset in every patient and tested differential abundance by Mann-Whitney U test across desired conditions. We then corrected for multiple hypotheses using Benjamini-Hochberg FDR for every lineage separately. We compared the different conditions we have in our single-cell cohort in two main levels: the first is comparing the finest resolution we have - pregnant-control / pregnant-asymptomatic / pregnant-severe / non-pregnant-asymptomatic / non-pregnant-severe. Since the sample size of these groups is limited, the majority of the comparisons came up as insignificant (**Figures S6E,K, S7C, S8B, S9D and S10G**). We, therefore, then tested differential abundance at a lower resolution by grouping all pregnant COVID-19 patient samples (pregnant-asymptomatic and pregnant-severe) and compared them to all non-pregnant COVID-19 patient samples (non-pregnant-asymptomatic and non-pregnant-severe) in which some cell subsets showed differential abundance (**Figures 3B** and **4H, Figures S7B, S8C** and **S10F**).

#### Repertoire analysis

##### Clone definitions, frequency of clones

**A clone** was defined by concatenating the following sequences that are the output of CellRanger for T cells: TRA_cdr3, TRB_cdr3, TRA_v_gene, TRB_v_gene, TRA_j_gene, TRB_j_gene; and for B cells: IGL_cdr3, IGK_cdr3, IGH_cdr3, IGL_v_gene, IGK_v_gene, IGH_v_gene, IGL_j_gene, IGK_j_gene, IGH_j_gene. Any cell that lacked any one of the above components was defined as having no clone information.

**Clone frequency** is defined as the ratio between the number of single cells with the exact same clone sequence for a patient and the total number of cells with TCR/BCR information for that patient (cells that have TCR/BCR information are illustrated in**, Figures S7D, S8D** and **S10A**).

An **expanded clone** is defined as a clone with a frequency of at least 1% out of a sample and that appears in at least 5 single cells in that sample.

#### Enrichment of TCR/ BCR clones in cell subsets

To identify enrichment of expanded clones in cell subsets in different conditions, we defined the frequency of expanded clones per cell subset per patient by

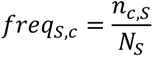

Where

*n_c,S_* is the total number of cells that are part of an expanded clone in sample S in cluster c

*N_S_* is the total number of cells that are part of an expanded clone in sample S

We then compared *freq_S,c_* across different conditions and used Mann-Whitney U test and Benjamini-Hochberg multiple hypothesis correction to test if they differ significantly (**Figure 3D, Figures S10F** and **G**).

#### Putative epitopes for CD8 TCRs

We downloaded the VDJdb^137^ database in February 2023 and queried it to identify putative epitopes for the CD8 TCRs identified in our data using an R script.

#### HLA alleles

In an attempt to make the putative epitopes found by VDJdb more specific (in the first query of VDJdb several epitopes were found for most TCRs), we sought to identify HLA alleles per patient and rerun the VDJdb query. To identify HLA alleles, we used the functions extract() and genotype() from the repo ArcasHLA^138^ (v0.5.0, R).

#### TCR sequence diversity

Diversity curves that measured Hill’s diversity metric across diversity orders 0–4 were created using the package alakazam^139^ (v1.2.1, R) with the alphaDiversity function. Hill’s diversity metric was only calculated on samples with ≥200 total TCR sequences. We tested the differences between the diversity index in q=[0,1,2,3,4] by a Mann-Whitney test.

#### Metagenomic analysis

Fresh stool samples were collected and refrigerated at 4 °C until aliquoting and freezing at -80 °C, typically within 4 hrs of collection. Nucleic acids were extracted from stool samples using the AllPrep PowerFecal DNA/RNA 96 Kit (Qiagen) and metagenomes were sequenced at the Broad Institute, according to their standard established platforms. Briefly, DNA was prepared for sequencing using the Illumina Nextera XT DNA library preparation kit and sequenced with a target of 3GB output at 2x150 bp read length using the NovaSeq platform (Illumina). Taxonomic profiles were generated using the bioBakery 3 shotgun metagenome workflow 3.0.0, which has previously been described^140^. Human reads were filtered using KneadData 0.10.0 and taxonomic profiles were generated using MetaPhlAn 3.0.0.

#### Mouse PR8 infection

Wild-type C57BL/6 mice were purchased from Taconic Biosciences. Female mice (8–10-weeks old) were anesthetized with an intraperitoneal injection of a mixture of 80 mg/kg ketamine and 16 mg/kg xylazine in PBS and infected with 50 PFU of PR8 intranasally. Body weight loss and survival were monitored. Survival was estimated according to the Kaplan–Meier method, and significance was calculated by the log-rank test. Mice were euthanized when they lost more than 20% of their initial body weight. For the IL-27 gain-of-function experiment, PR8-infected mice were intraperitoneally injected daily with 500 ng of recombinant mouse IL-27 (BioLegend) or PBS from day 0 to day 7. PR8 infectivity and lung damage-associated marker genes were measured by qPCR. Lung tissues were stored in RNAlater solution at -80 °C until further processing. RNA was extracted by TRIzol (Invitrogen) following the manufacturer’s instructions. RNA concentration was quantified and normalized prior to cDNA synthesis. cDNA was synthesized by reverse transcription (iScript, Bio-Rad), followed by qRT-PCR (LightCycler 96, Roche Life Science) using SYBR green (iTaq universal SYBR, Bio-Rad) and listed primers. PCR results were normalized against beta-actin.

The following gene-specific forward (F) and reverse (R) primers were used.

Beta-actin F: 5’-GGCTGTATTCCCCTCCATCG-3’ R: 5’-CCAGTTGGTAACAATGCCATGT-3’; Muc5ac F: 5’-TACCACTCCCTGCTTCTGCAGCGTGTCA-3’ R: 5’-ATAGTAACAGTGGCCATCAAGGTCTGTCT-3’; F3 F: 5’-GTGCTTCTCGACCACAGACA-5’ R: 5’-GGCTCCTCCCCATGAATCAC-3’; Retnla F: 5’-CCCTTCTCATCTGCATCTCCC-3’ R: 5’-AAGCACACCCAGTAGCAGTC-3’; Ns1 F: 5’-CTGTGTCAAGCTTTCAGGTAGA-3’ R: 5’-GGTACAGAGGCCATGGTCAT-3’; Np F: 5’-CCCAGGATGTGCTCTCTGAT-3’ R: 5’-TTCGTCCATTCTCACCCCTC-3’

#### Albumin detection in BAL fluids

To detect albumin leakage in BAL, PR8-infected mice were euthanized 3 days post-infection. The thorax of each mouse was then opened, and a 20 GA × 1.16 IN (1.1 mm × 30 mm) I.V. catheter (BD) was introduced into the trachea at the cricothyroid membrane. The trachea was then flushed with 1000 µl PBS, and the resulting fluid was aspirated. Collected BAL fluids were stored at −80 °C. Albumin levels in BALs were measured with the mouse albumin ELISA kit (Abcam)

#### IL-27 detection in mouse models

To monitor IL-27 expression in the anti-CD3 injection-induced inflammation model, mice were intraperitoneally injected with 20 µg of anti-CD3 (Tonbo Bioscience). At day 3, mice were euthanized, and blood was collected via cardiac puncture. To monitor IL-27 expression in non-pregnant, syngenic (B6 x B6), or allogenic (CBA x B6) pregnant mice, male B6 or CBA background mice were mated overnight with 8–10-week-old B6 females. At E17.5, mice were anesthetized, and blood was collected with retro-orbital bleeding. For plasma isolation, all blood samples were centrifuged at 10,000 × *g* for 10 min at 4 °C. IL-27 levels were monitored with an ELISA kit (R&D systems).

#### Fetal demise experiment

To induce fetal demise in pregnant mice, E12.5 pregnant mice were injected with 600 µg poly(I:C) (HMW) VacciGrade (InvivoGen). At E17.5, mice were euthanized, and the fetuses were collected for evaluation. The total number of fetuses and the percentage of pups scored nonviable or resorbed fetuses were calculated.

#### Quantification and statistical analysis

Statistical analyses were performed with PRISM 9. Each data point denotes individual human subjects or animals. For **Figures 1–2, 3, S2–S5, S11, S14 and S15** statistical significance was calculated by Mann-Whitney non-parametric test for comparisons between the non-pregnant and pregnant groups in the same disease severity, and Kruskal-Wallis non-parametric test for comparisons within the non-pregnant or pregnant group. *P-values* were shown after multiple hypotheses correction by Dunn’s. For the boxplots in **Figures 3, 4,** and **S6-10**, each dot represents a patient, the box limits represent the 25^th^ and 75^th^ percentiles of the data distribution (inter-quartile range, IQR), the line in the middle of the box represents the median and the whiskers represent data points up to 1.5*IQR. Dots beyond the whiskers are considered outliers and are marked by a diamond. Statistical testing - Mann-Whitney U test; *P-values* were shown after multiple hypotheses correction by Benjamini-Hochberg; (ns) denotes not significant after multiple hypotheses correction. For **Figure 6**, statistical significance was calculated by two-way ANOVA for (C). A *P-value* for survival rate was calculated by the Gehan-Breslow-Wilcoxon test for (D). *P-values* for (E), (F), (G), and (I) were calculated by unpaired t-test.

## Contributions

A.G.E. and J.R.H. conceptualized the study. D.S., E.K., G.L., G.B.C., A.G.E., and J.R.H. designed experiments. D.S., E.K., G.L., R.N., A.L., and O.P. analyzed data. MGH COVID Collection & Processing Team, P.S., J.T., A.T., B.Y.A., collected, processed and generated samples and/or data from the MGH acute COVID19 non-pregnant cohort, which was overseen by X.Y. and J.Z.L. L.L.S., O.J., S.D., E.G., J.K. and B.A. enrolled, collected, processed and generated samples and phenotypic data from the MGB COVID-19 Pregnancy Biorepository, which was overseen by A.G.E. and K.J.G., and to which L.M.Y. and A.F. also contributed personnel support and resources. M.B.G., M.R.F, and N.H. oversaw the MGH acute COVID cohort. P.S., J.T., A.T., and B.Y.A. performed the single-cell multiomics experiments for the COVID19 pregnancy cohort, R.N. performed single cell data analysis. R.N., P.S., and A-C.V performed single cell data analysis interpretation. A-C.V. managed and supervised scRNAseq data generation and analysis. D.A.D. and L.H.N. performed metagenomic sequencing and analysis, supervised by A.C.. D.S., E.K., G.L., R.N., P.S., A-C.V., A.G.E., and J.R.H. wrote the manuscript. All authors read or provided comments on the manuscript.

## Below is the list of all MGH COVID Collection & Processing Team (Non-Pregnant Samples) Participants

### Collection Team

Kendall Lavin-Parsons, Blair Parry, Brendan Lilley, Carl Lodenstein, Brenna McKaig, Nicole Charland, Hargun Khanna, Justin Margolin

Department of Emergency Medicine, Massachusetts General Hospital, Boston, MA 02115 Processing Team:

Anna Gonye, Irena Gushterova, Tom Lasalle, Nihaarika Sharma Massachusetts General Hospital Cancer Center, Boston, MA 02115

Brian C. Russo, Maricarmen Rojas-Lopez

Division of Infectious Diseases, Department of Medicine, Massachusetts General Hospital, Boston, MA 02115

Moshe Sade-Feldman, Kasidet Manakongtreecheep, Jessica Tantivit, Molly Fisher Thomas, Thomas Lasalle, Thomas Eisenhaure

Massachusetts General Hospital Center for Immunology and Inflammatory Diseases, Boston, MA 02115

## Data and code availability

scRNA data, CITE-seq, and repertoire data were deposited to GEO and are available in the accession number GSE239452. The code used for single-cell analysis and figures generation will be available in GitHub at https://github.com/villani-lab/COVID_pregnancy_and in the Zenodo repository upon publication of this study. For reviewers, we have created a compressed file that contains all the code as part of our submission documents (COVID_pregnanacy_code.zip). A full list of software packages and versions included in the analysis can be found in Supplementary Table 10.

## Supplementary Figures

**Figure S1.**
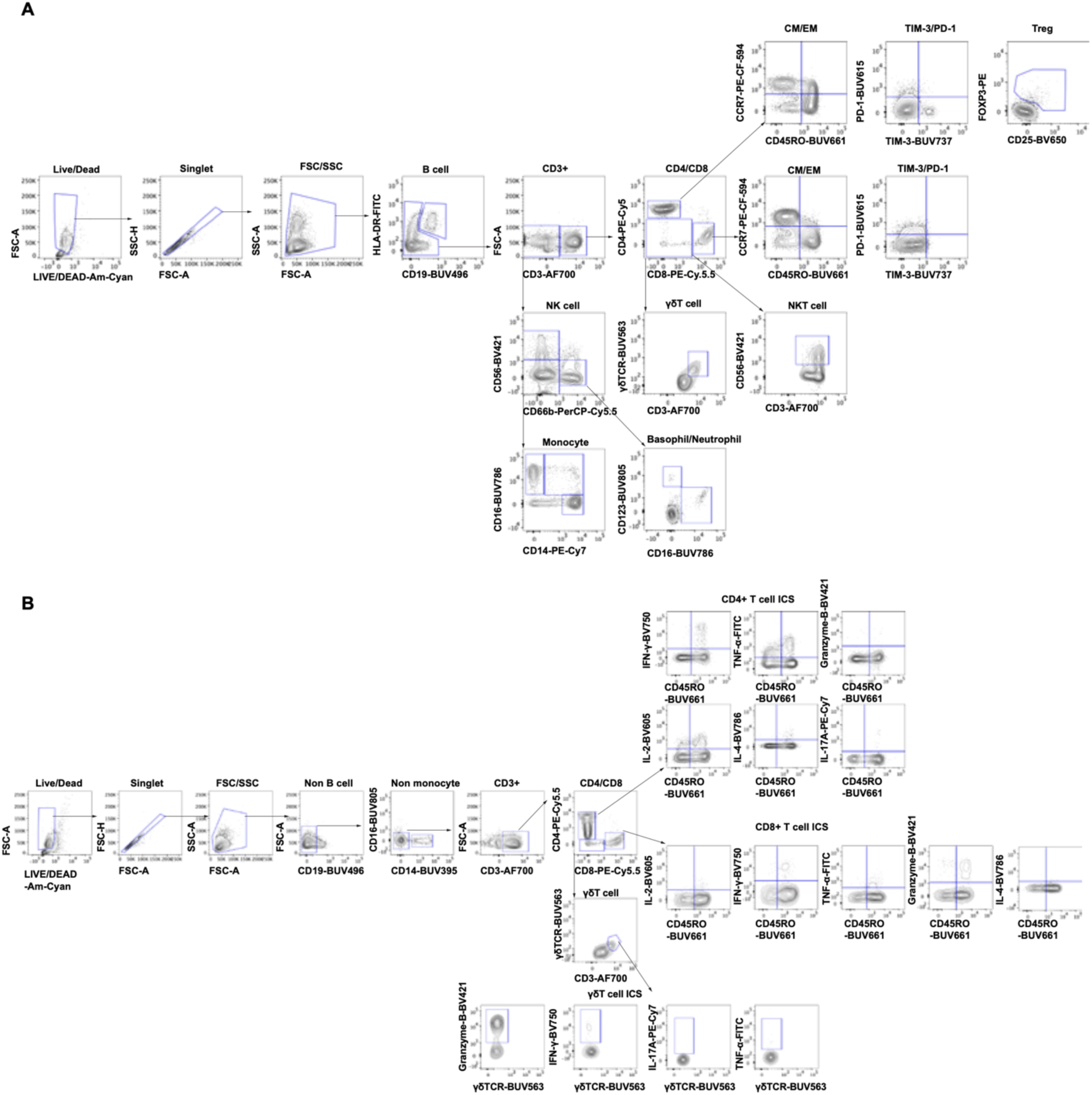
Gating strategies for flow cytometry analysis. (**A**) Gating strategy for defining immune cell populations. Dead cells were excluded by LIVE/DEAD™ Fixable aqua dead cell staining. Doublet and cell debris were removed by singlet gating and FSC SSC-based gating. CM and EM T cells were defined by CD45RO and CCR7 expression. (**B**) Gating strategy for analyzing intracellular cytokine production from CD4^+^, CD8^+^, and γδ-T cells.

**Figure S2.**
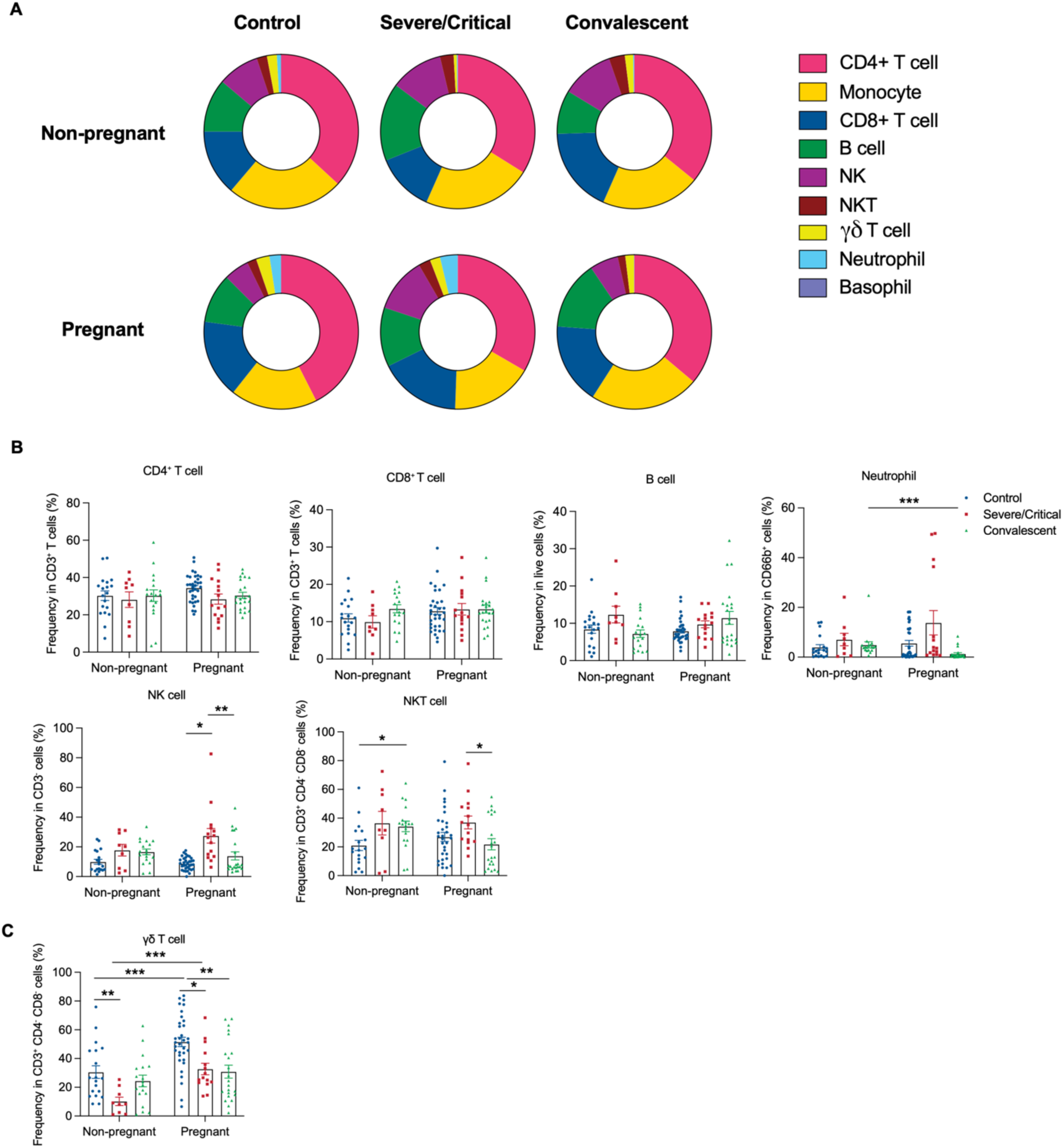
Immune cell frequencies in PBMCs. (**A**) Pie chart of immune cell populations in PBMCs. The average frequency of each cell type among total live cells is displayed. (**B**) Frequencies of T (CD19^-^ CD3^+^ CD4^+^ or CD8^+^) and B (CD19^+^ HLA-DR^+^), neutrophil (CD19^-^ CD3^-^ CD66b^+^ CD16^+^), NK (CD19^-^ CD3^-^ CD56^+^ CD66b^-^) and NKT (CD19^-^ CD3^+^ CD4^-^ CD8^-^ CD56^+^) cells in PBMCs. (**C**) Frequencies of γ8T cells (CD19^-^ CD3^+^ CD4^-^ CD8^-^γ8 TCR^+^) in PBMCs. Healthy (Non-pregnant n = 19, Pregnant n = 34) or who had Severe/Critical COVID-19 (Non-pregnant n = 9, Pregnant n = 15) or convalescent COVID-19 (Non-pregnant n = 18, Pregnant n = 22). Data are shown as the mean ± SEM. *p < 0.05, **p < 0.01, ***p < 0.001.

**Figure S3.**
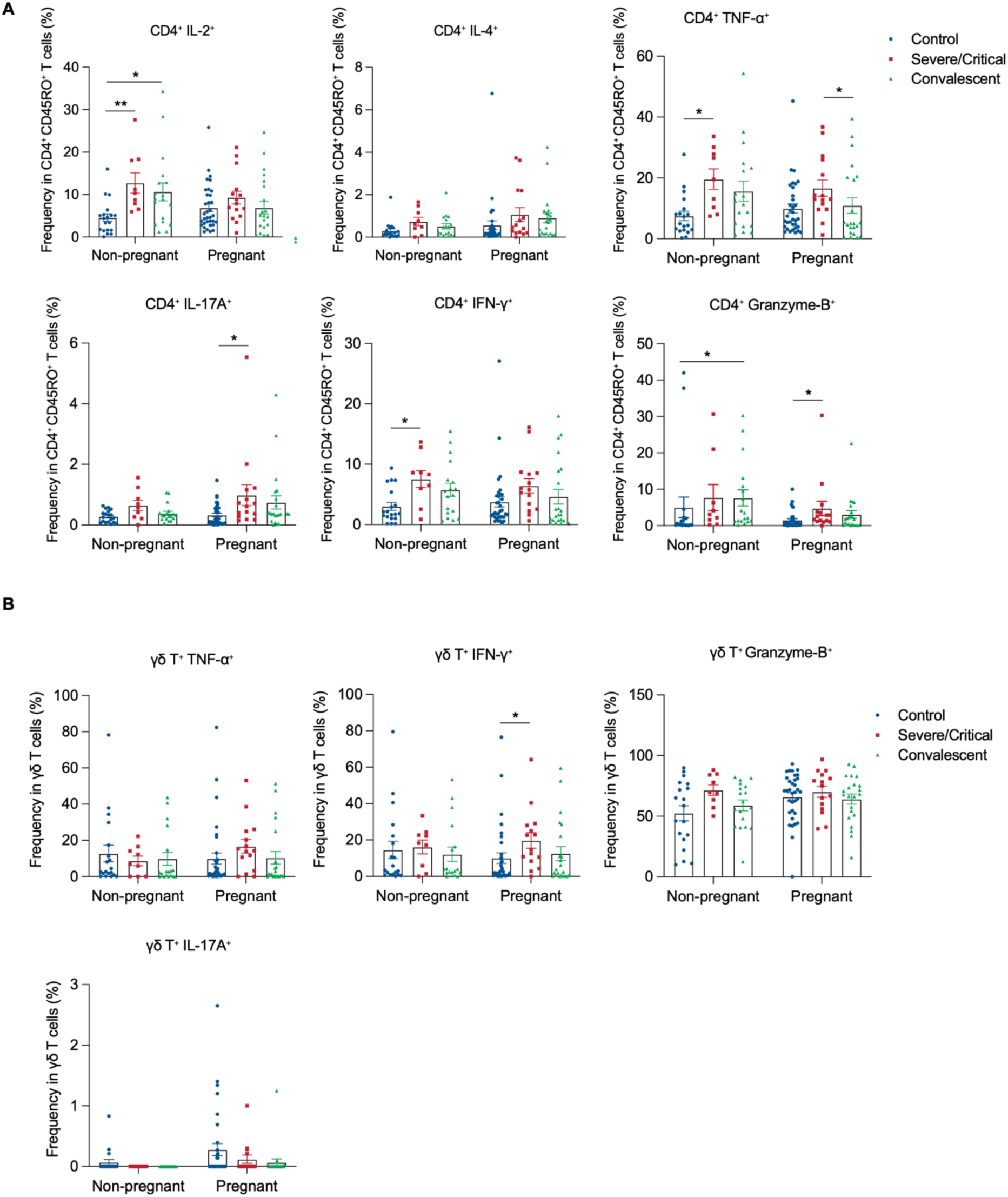
Cytokine-producing CD4^+^ T cells and γδT cells in COVID-19 women. (**A** and **B**) Frequencies of cytokine-producing CD4^+^ T cells (**A**) and γδ-T cells (**B**) in PBMCs from healthy (Non-pregnant n = 19, Pregnant n = 34) or who had Severe/Critical COVID-19 (Non-pregnant n = 9, Pregnant n = 15) or convalescent COVID-19 (Non-pregnant n = 18, Pregnant n = 22). Data are shown as the mean ± SEM. *p < 0.05, **p < 0.01.

**Figure S4.**
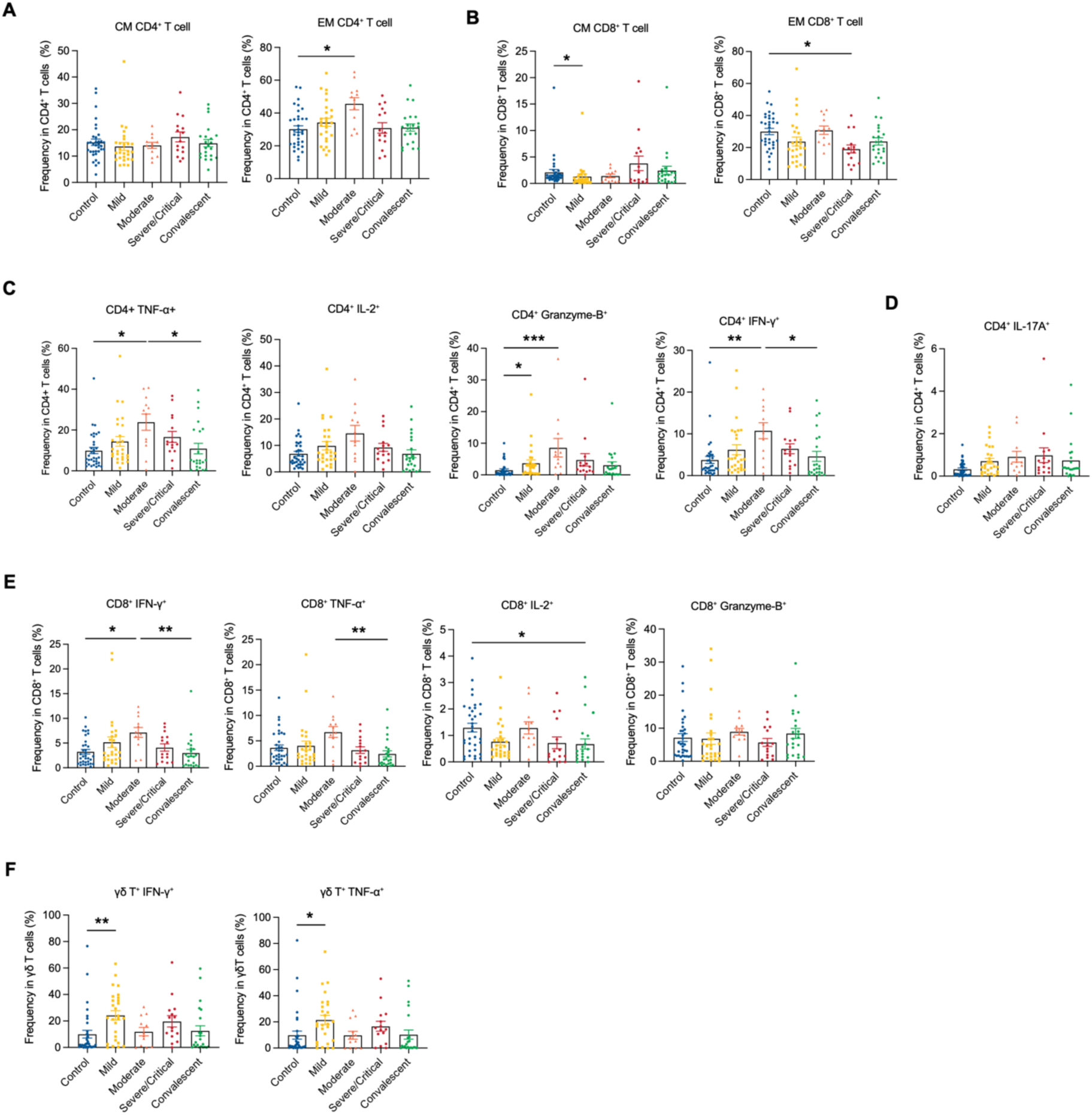
Comparison of cytokine production in T cells from pregnant patients with different disease severity in COVID-19. (**A** and **B**) CM and EM frequencies of CD4^+^ and CD8^+^ T cells in PBMCs from. (**C** to **E**) Frequencies of cytokine-producing CD4^+^ (**C**), IL-17A^+^ producing CD4^+^ (**D**), and CD8^+^ (**E**) T cells in PBMCs from pregnant subjects. (**F**) Frequencies of cytokine-producing γδ-T cells in PBMCs from pregnant subjects. Healthy pregnant (n = 34) or who had mild (n = 27) or moderate (n = 12) or severe/critical (n = 15) or convalescent COVID-19 (n = 22) in pregnant women. Data are shown as the mean ± SEM. *p < 0.05, **p < 0.01, ***p < 0.001.

**Figure S5.**
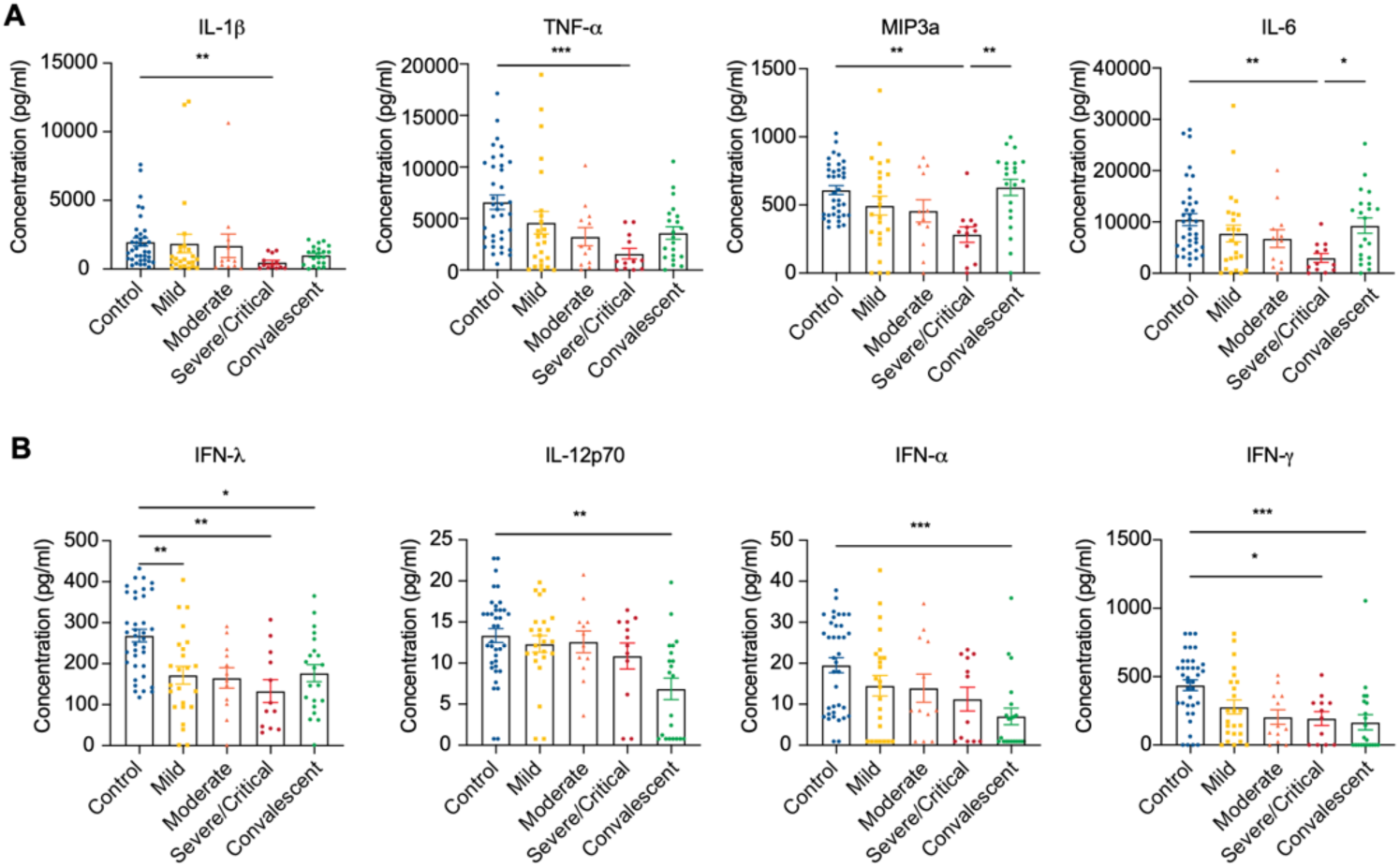
Monocyte-secreted cytokines depend on the disease severity within the pregnancy. (**A** and **B**) Cytokine production from CD14^+^ monocytes from upon LPS stimulation was compared within pregnancy groups (healthy pregnant n = 37) or who had mild (n = 24) or moderate (n = 12) or severe/critical (n = 12) or convalescent COVID-19 (n = 21) in pregnant women. Data are shown as the mean ± SEM. *p < 0.05, **p < 0.01, ***p < 0.001.

**Figure S6.**
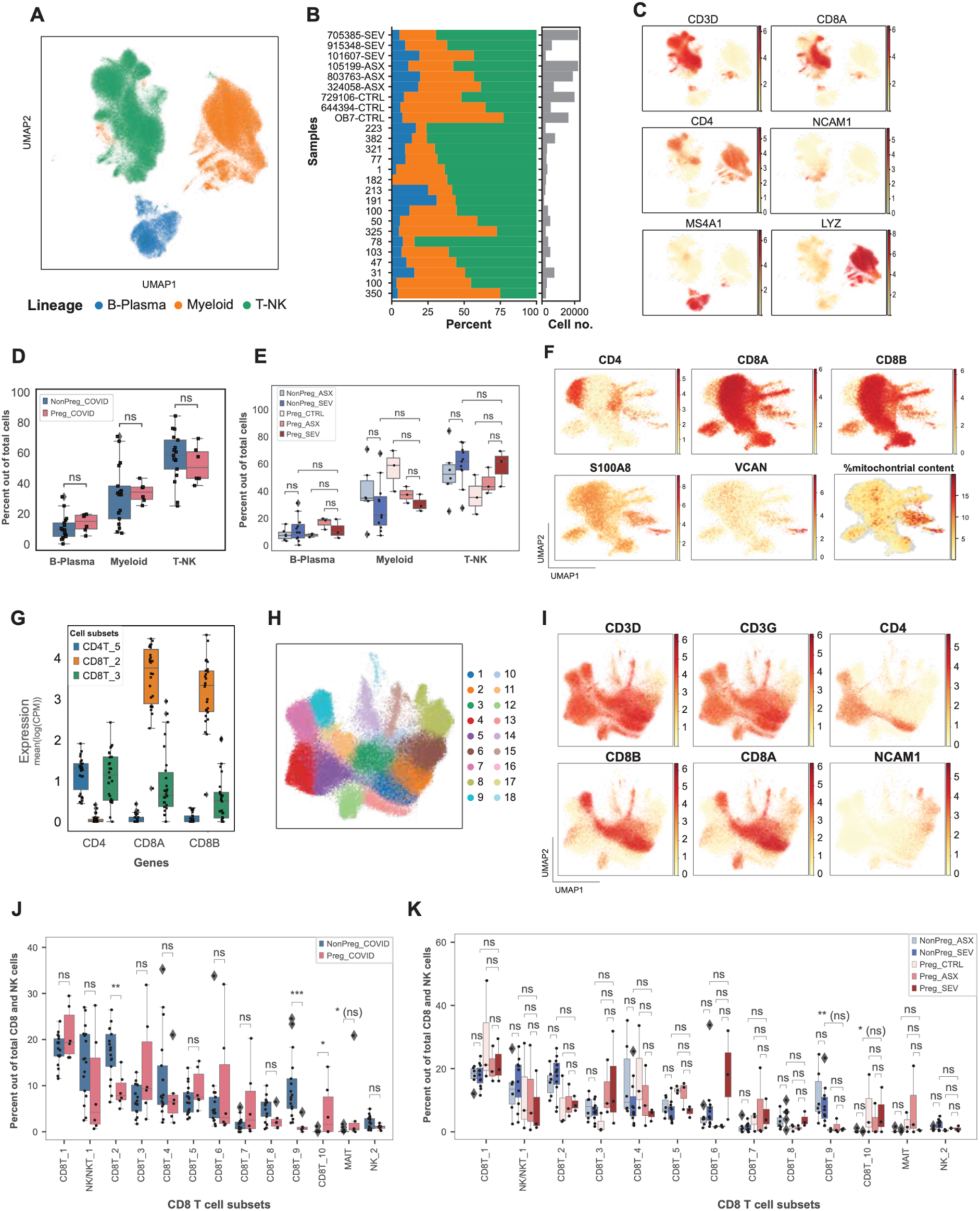
Global single-cell clustering and CD8^+^ T cell clustering and differential abundance. (**A**) UMAP embedding of all 181,112 single cells in the cohort colored by main lineages. (**B**) Percentage of cells in each lineage per patient (left) and total number of cells per patient (right). (**C**) Expression of marker genes that characterize the main lineages, log(CPM). (**D**) A boxplot illustrating the abundances of the different cell lineages in pregnant vs. non-pregnant patients with COVID-19. (**E**) A boxplot illustrating the abundances of the different cell lineages in pregnant vs. non-pregnant individuals and across COVID-19 severities. (**F**) Gene expression was used to identify doublets and cell subsets with high mitochondrial content. (**G**) A boxplot illustrating the gene expression of CD4, CD8A, and CD8B in CD8T_3 compared to a “pure CD8” T cell subset (CD8T_2) and a “pure CD4” T cell subset (CD4T_5) demonstrating CD8 is expressed in low levels in CD8T_2, and CD4 is expressed at high levels in this specific cell subset. (**H**) UMAP of all CD4 T, CD8 T, and NK cells. (**I**) Gene marker expression was used to split the cells in Figure S6H into CD4 and CD8 T cell sub-lineages, log(CPM). (**J**) A boxplot illustrating the abundances of CD8 and NK cell subsets in pregnant vs. non-pregnant patients with COVID-19. (**K**) A boxplot illustrating the abundances of CD8 and NK cell subsets in pregnant vs. non-pregnant patients with COVID-19 and across COVID-19 severities. *p < 0.05, **p < 0.01, ***p < 0.001, p-values are shown after multiple hypothesis correction, if any test becomes not significant after multiple hypothesis correction it is denoted with ‘(ns)’ next to the un-adjusted p-value.

**Figure S7.**
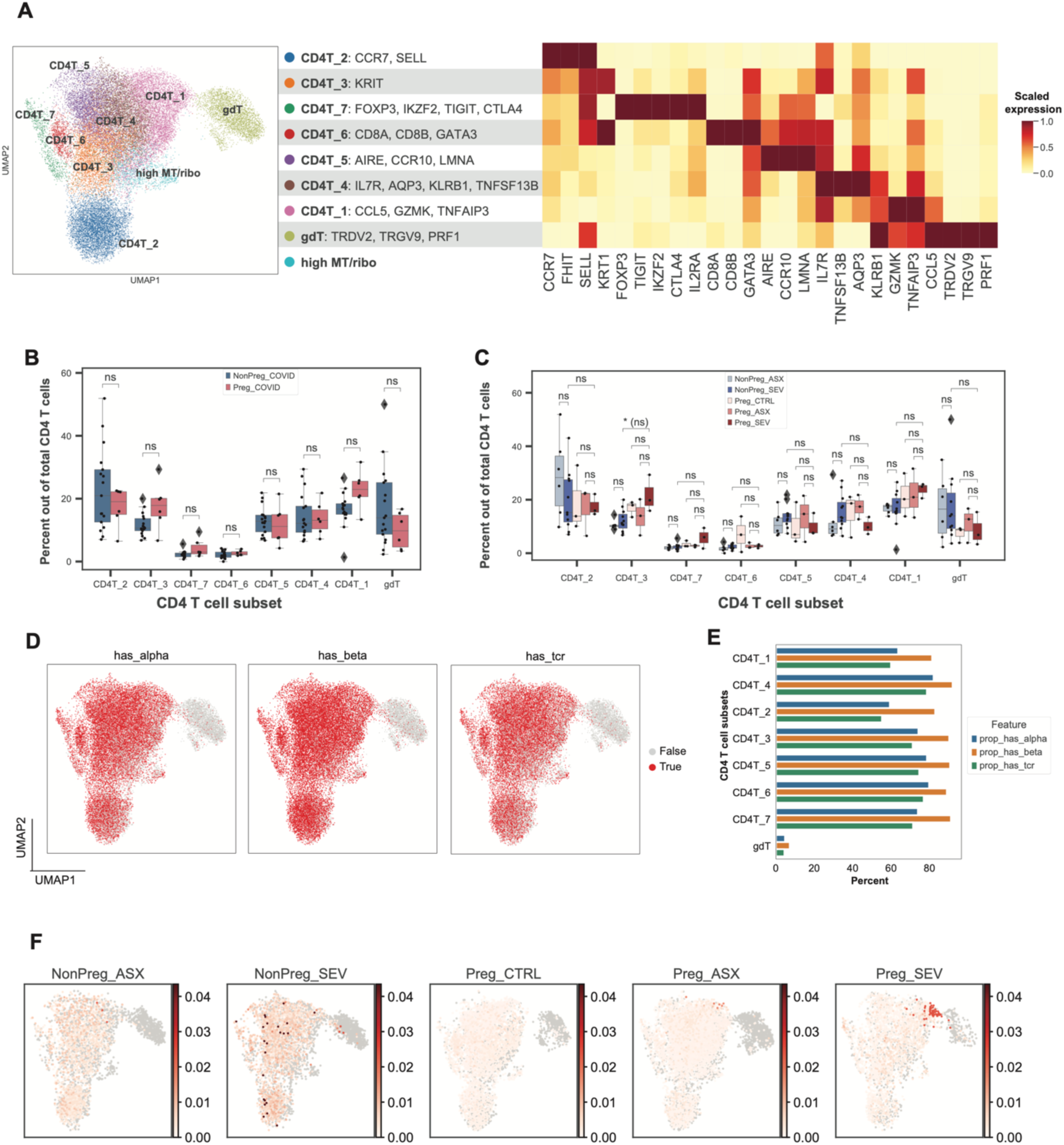
CD4^+^ T cell clustering, cell subset abundance, and clonal expansion information. (**A**) UMAP embedding of 29,960 CD4^+^ T cells and their clustering to eight cell subsets (left). Expression of main marker genes, scaled mean log(CPM) (right). (**B**) A boxplot illustrating the abundance of CD4^+^ T cell subsets in pregnant vs. non-pregnant COVID-19 patients. (**C**) A boxplot illustrating the abundance of the CD4^+^ T cell subsets in pregnant vs. non-pregnant individuals and across COVID-19 severities. (**D**) UMAP projection of cells that have alpha chain sequence, beta chain sequence, and both sequences with the V and J genes (has_tcr) highlighting cell subsets with TCR data. (**E**) Quantification of features shown in Figure S5E per cell subset. (**F**) Clone frequency per patient is shown on the CD4^+^ T cells UMAP. *p < 0.05, **p < 0.01, ***p < 0.001, p-values are shown after multiple hypothesis correction, if any test becomes not significant after multiple hypothesis correction it is denoted with ‘(ns)’ next to the un-adjusted p-value.

**Figure S8.**
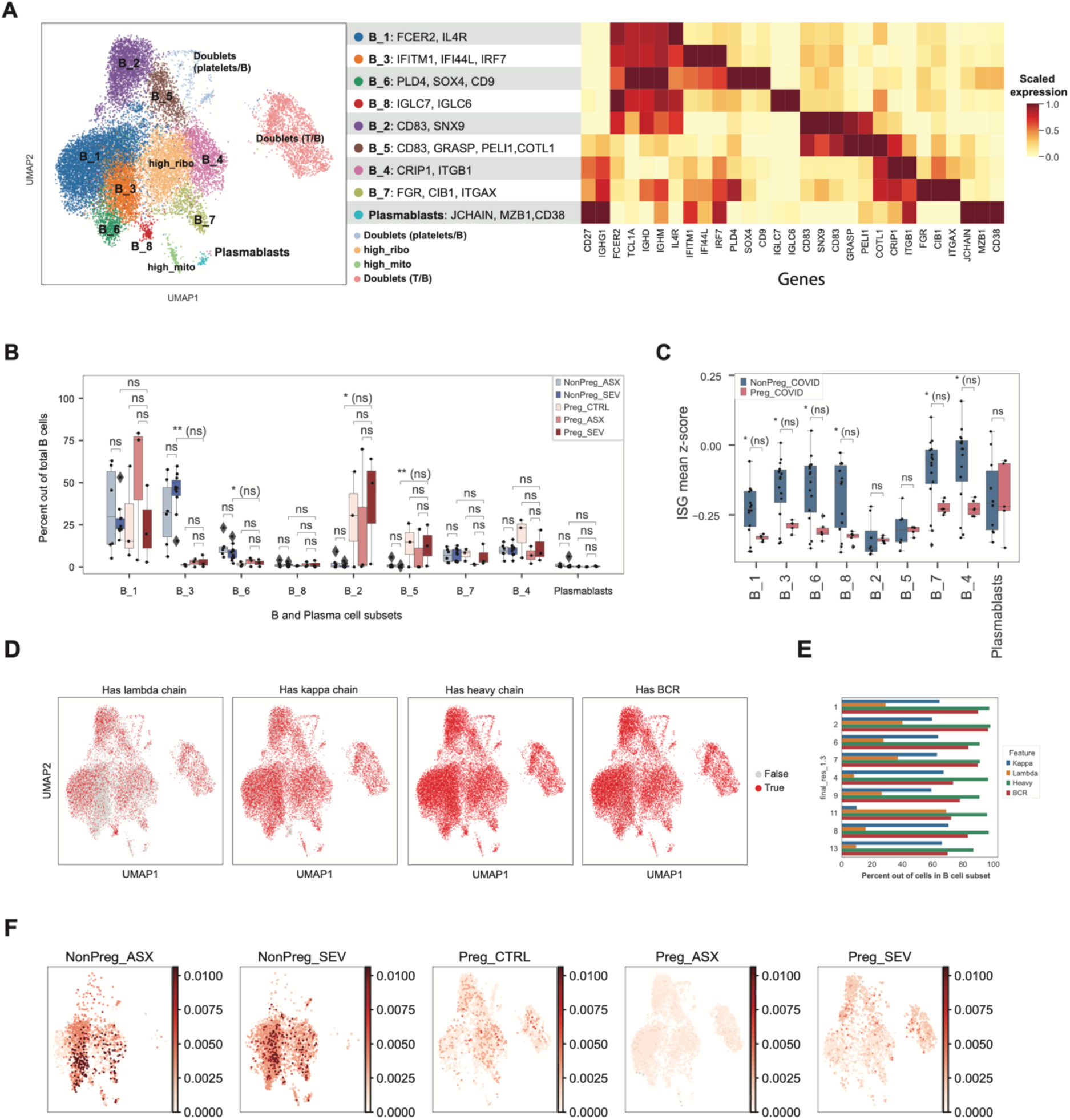
B cell clustering, cell subset abundance, and clonal expansion information. (**A**) UMAP embedding of 19,168 B and plasma cells and their clustering to nine cell subsets (left). Expression of main marker genes, scaled mean log(CPM) (right). (**B**) A boxplot illustrating the abundances of the different B cell subsets in pregnant vs. non-pregnant individuals and across COVID-19 severities. (**C**) A boxplot illustrating the ISG expression signature in different B cell subsets in pregnant vs. non-pregnant COVID-19 patients. (**D**) UMAP projection of cells that have the lambda chain sequence, kappa chain sequence, heavy chain sequence, and either kappa or lambda *and* a heavy chain with the V and J genes (“has bcr”) from the repertoire sequencing, highlighting cell subsets with BCR data. (**E**) Quantification of features shown in Figure S9D per cell subset. (**F**) Frequency of BCR clones projected on B cell UMAP and split by condition (pregnant and non-pregnant; severity of COVID-19 infection). *p < 0.05, **p < 0.01; p-values are shown after multiple hypothesis correction, if any test becomes not significant after multiple hypothesis correction it is denoted with ‘(ns)’ next to the un-adjusted p-value.

**Figure S9.**
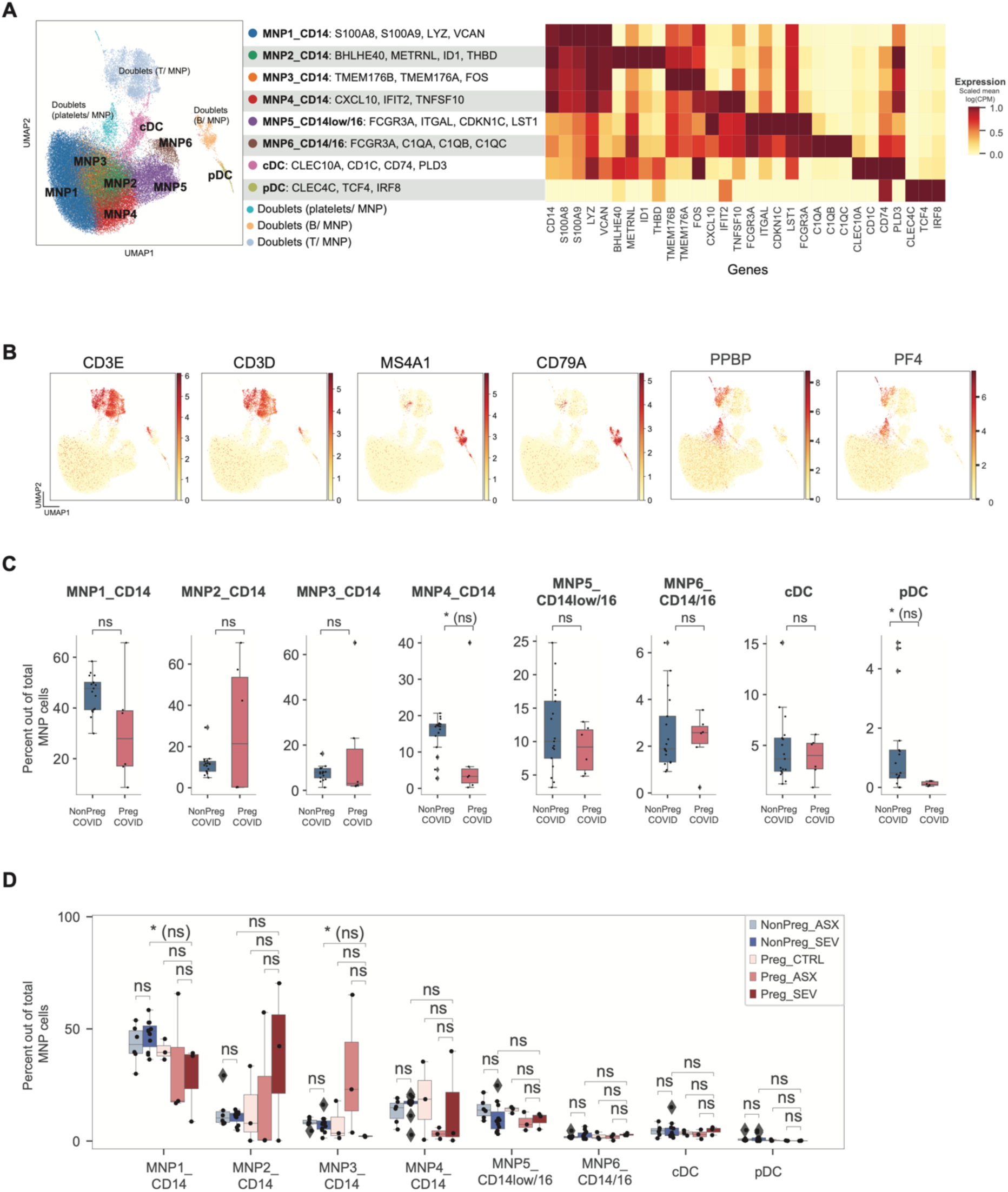
MNP cell clustering and cell subset abundance. (**A**) UMAP embedding of 66,719 mononuclear phagocyte (MNP) cells and their clustering to eight cell subsets (left). Expression of main marker genes, scaled mean log(CPM) (right). (**B**) Gene expression of main markers used to identify doublet populations, scaled mean log(CPM). (**C**) Boxplots illustrating the abundance of MNP cell subsets out of all MNP cells in pregnant COVID-19 patients vs. non-pregnant COVID-19 patients. (**D**) A boxplot illustrating the abundances of the different MNP cell subsets in pregnant vs. non-pregnant individuals and across COVID-19 severities. *p < 0.05; ‘ns’ denotes not significant; p-values are shown after multiple hypothesis correction, if any test becomes not significant after multiple hypothesis correction it is denoted with ‘(ns)’ next to the un-adjusted p-value.

**Figure S10.**
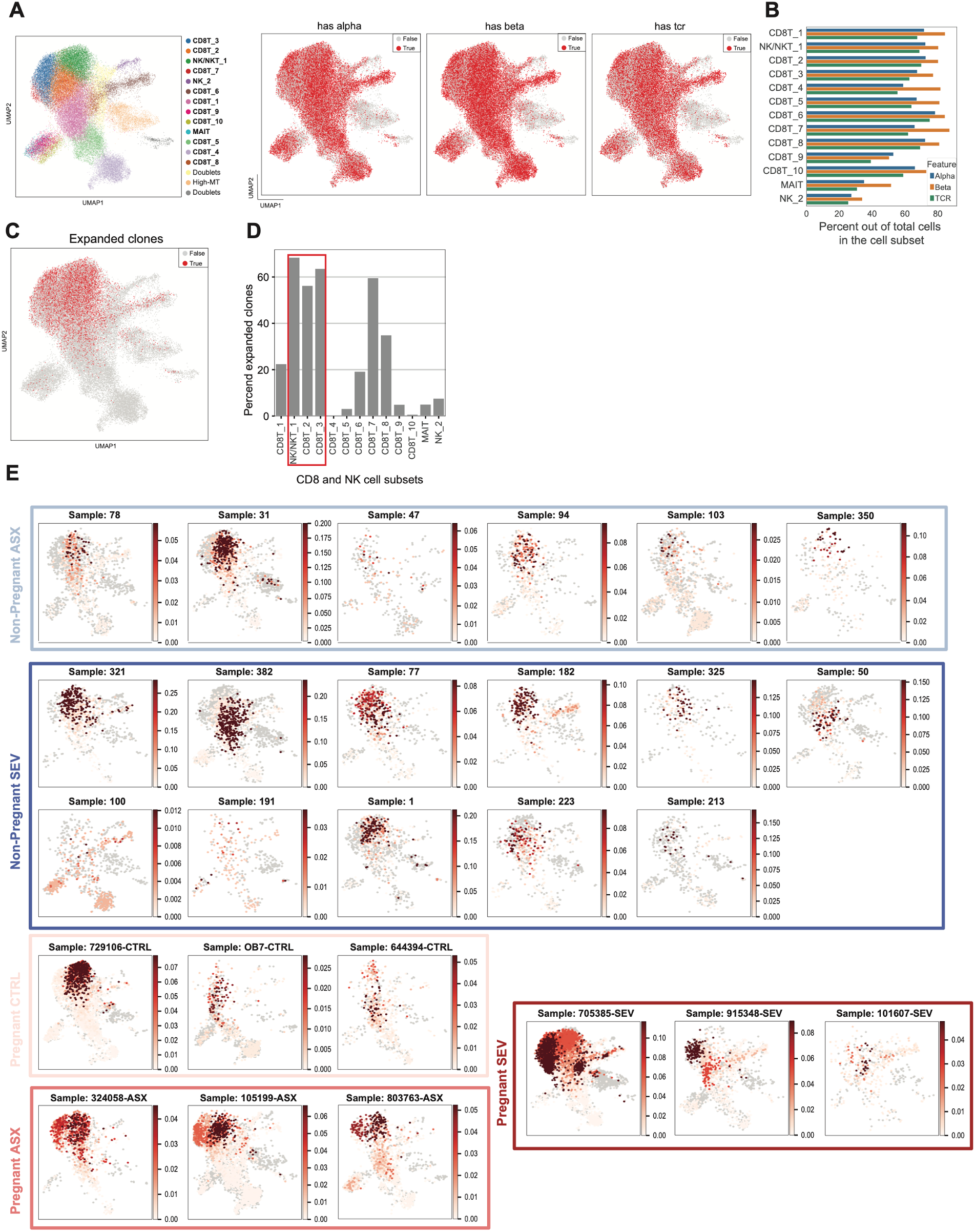
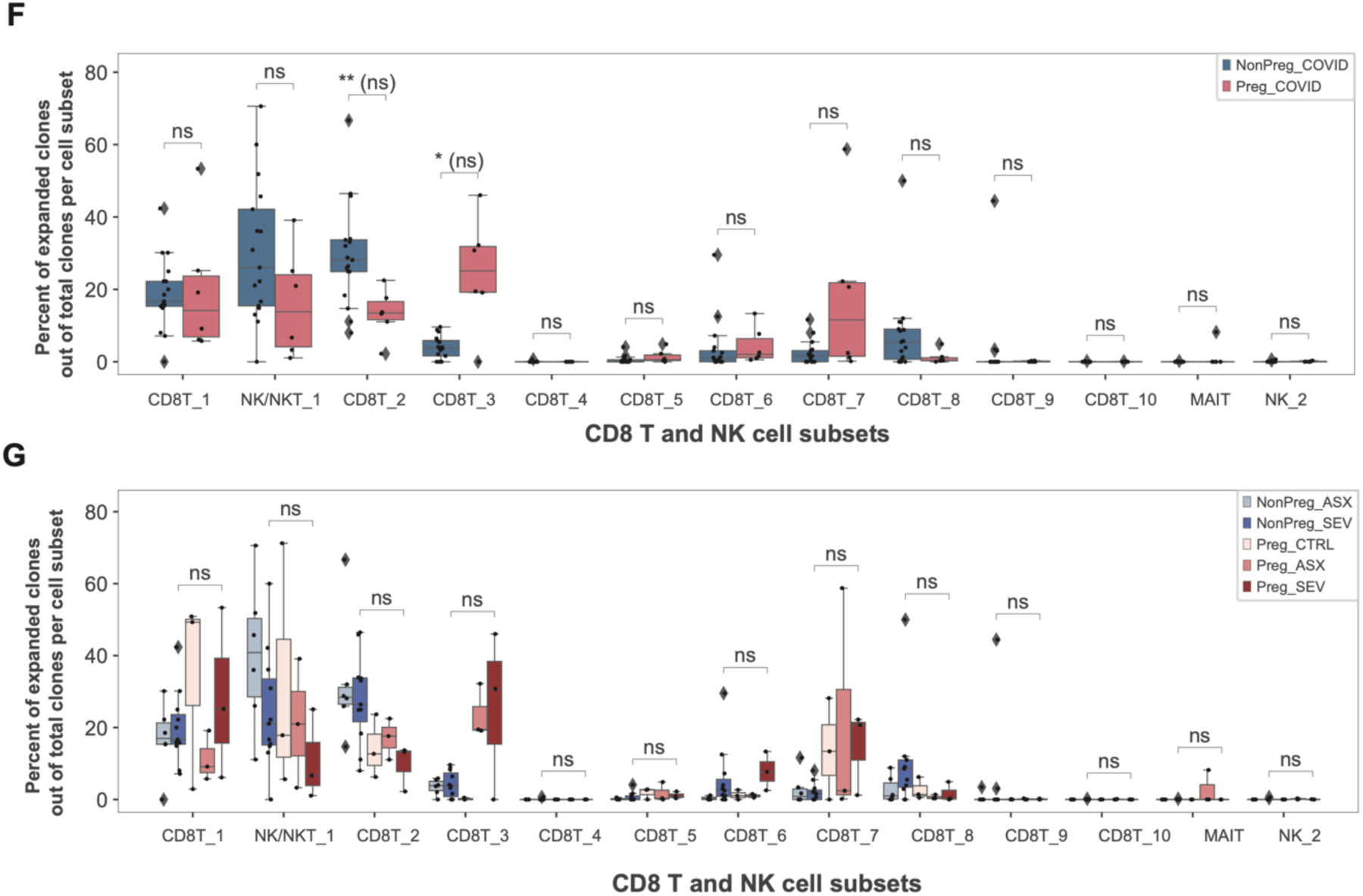
CD8^+^ T cell repertoire data information and clonal expansion across clusters and donors. **(A)** UMAP projection of cells with alpha chain sequence, beta chain sequence, and sequences with the V and J genes (has_tcr), highlighting cell subsets with TCR data. (**B**) Quantification of features shown in Figure S7A per cell subset. (**C**) Expanded clones are shown on the UMAP of CD8^+^ T and NK cells. (**D**) Quantification of Figure S7C shows the percentage of expanded clonal cells out of all cells in each cell subset with TCR data. (**E**) Clone frequency per patient is shown on the CD8^+^ T and NK UMAP. (**F**) A boxplot illustrating the percent of expanded clones out of total clones per cell subset, shown for pregnant and non-pregnant COVID-19 patients. (**G**) A boxplot illustrating the percent of expanded clones out of total clones per cell subset, shown for pregnant and non-pregnant individuals and across COVID-19 severities. *p < 0.05, **p < 0.01; p-values are shown after multiple hypothesis correction, if any test becomes not significant after multiple hypothesis correction it is denoted with ‘(ns)’ next to the un-adjusted p-value.

**Figure S11.**
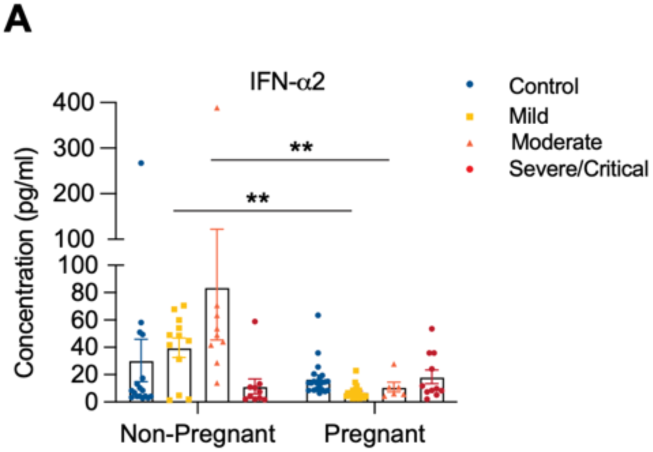
Decreased IFN-ɑ2 expression in pregnant COVID-19 subjects. Plasma IFN-ɑ2 was measured in pregnant COVID-19 samples from healthy (Non-pregnant n = 17, Pregnant n = 21) or who had Mild (Non-pregnant n = 12, Pregnant n = 21), Moderate (Non-pregnant n = 9, Pregnant n = 6), or Severe/Critical (Non-pregnant n = 10, Pregnant n = 11) COVID-19. Data are shown as the mean ± SEM. **p < 0.01.

**Figure S12.**
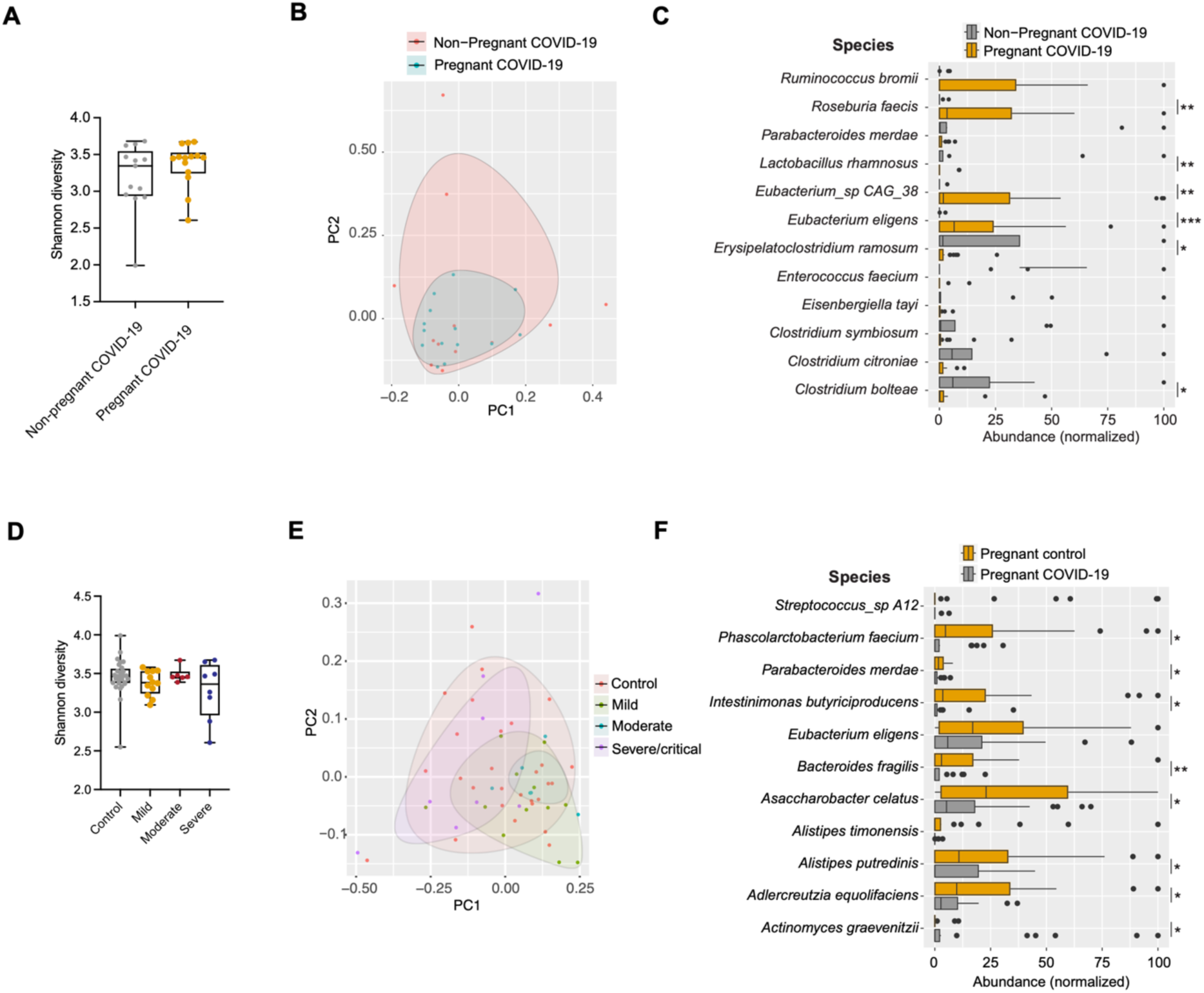
Microbiome analyses in stool samples from non-pregnant and pregnant subjects with COVID-19. (**A**–**C**) Metagenomic sequencing data from the fecal samples of non-pregnant and pregnant women with moderate (non-pregnant n = 4, pregnant n = 6) or severe COVID-19 (non-pregnant n = 9, pregnant n = 8) infection. (**A**) Shannon diversity of fecal microbiota based on metagenomic sequencing. (**B**) Beta-diversity (Bray-Curtis distance) between groups. (**C**) Species-level taxa based on relative abundance showing statistically significant differences. (**D**–**F**) Data of fecal samples collected from pregnant women with different clinical severities of COVID-19 (Healthy control n = 29, mild n = 13, moderate n = 6 and severe/critical n = 8). (**E**) Shannon diversity of fecal microbiota based on metagenomic sequencing. (**F**) Beta-diversity (Bray-Curtis distance) among groups. (**G**) Species-level taxa based on relative abundance show statistically significant differences. *p < 0.05, **p < 0.01, ***p < 0.001.

**Figure S13.**
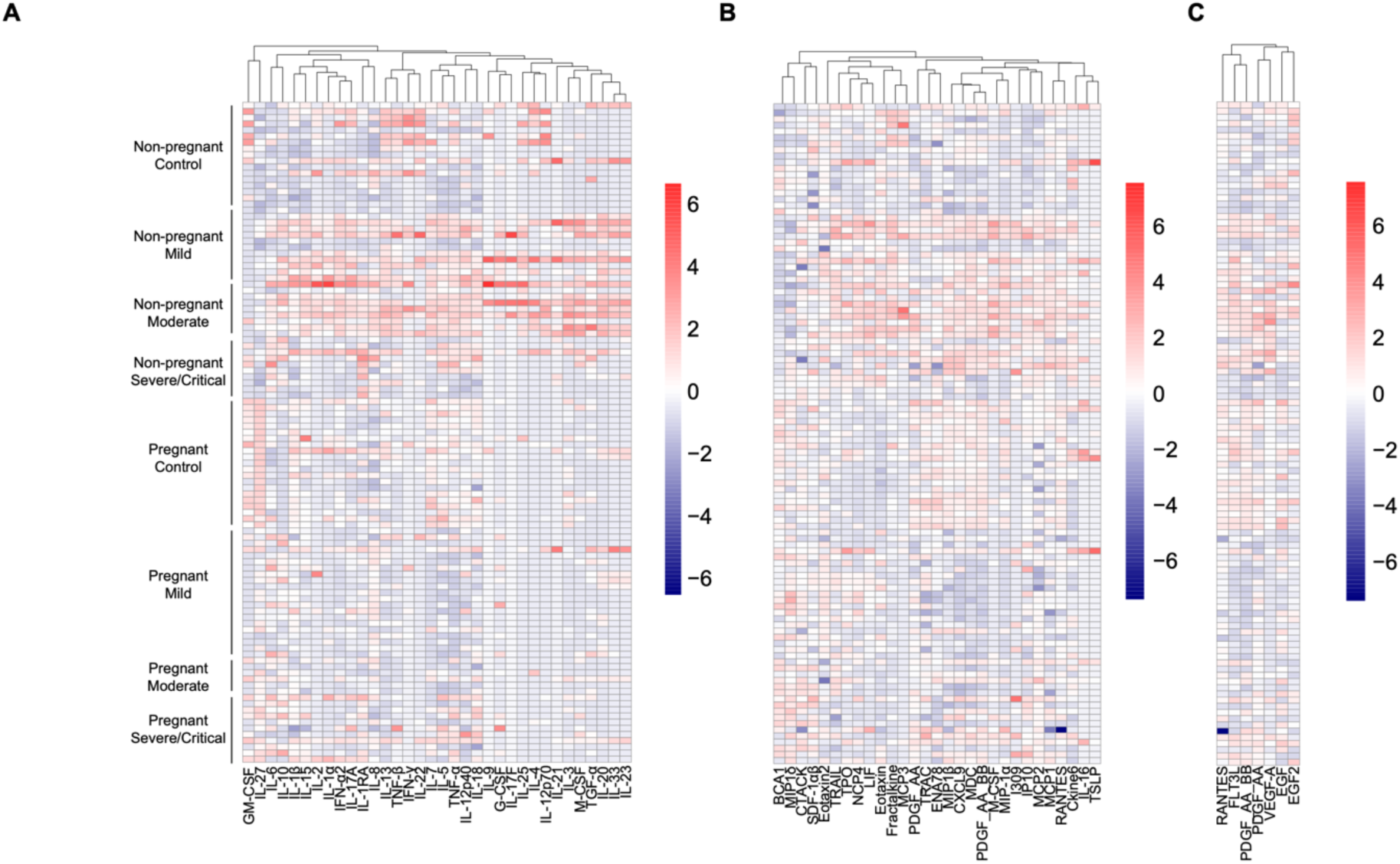
Heatmaps presenting cytokine, chemokine, and growth factor levels in the plasma of patients with COVID-19 and controls. Overview of cytokines (**A**), chemokines (**B**), and growth factors (**C**) in the plasma of both pregnant and non-pregnant healthy women and COVID-19 patients. Measurements were normalized across all samples.

**Figure S14.**
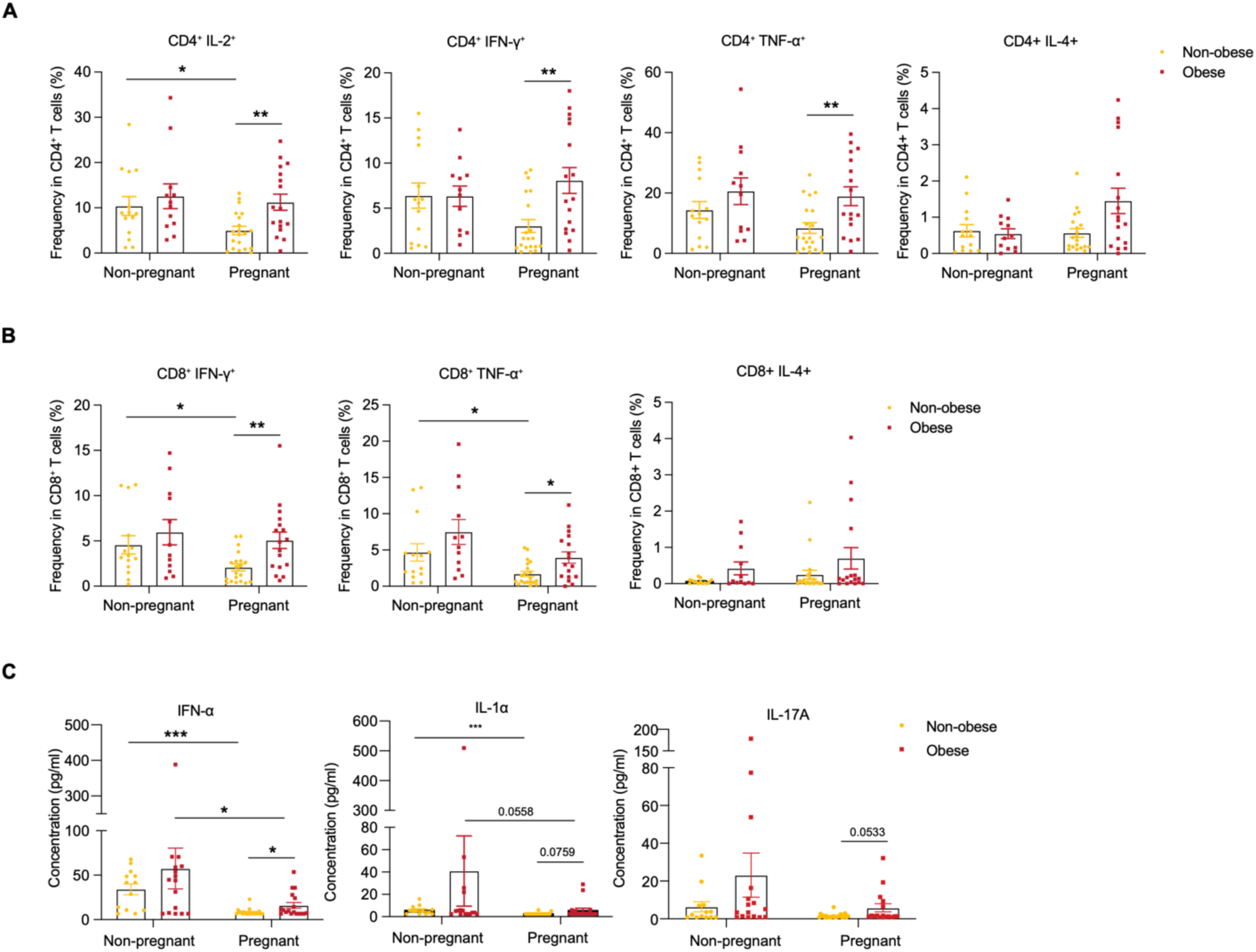
Pre-pregnancy BMI is associated with increased T cell production of pro-inflammatory cytokines in COVID-19. (**A**) Cytokine production in CD4^+^ T cells and (**B**) CD8^+^ T cells in PBMCs from non-obese COVID-19 (Non-pregnant n = 14, Pregnant n = 20) or obese COVID-19 (Non-pregnant n = 12, Pregnant n = 17) measured by flow cytometry. (**C**) Plasma from non-obese COVID-19 (Non-pregnant n = 13, Pregnant n = 22) or obese COVID-19 (Non-pregnant n = 16, Pregnant n = 16) cytokine levels measured by multiplex-based assay. All samples were from non-pregnant and pregnant patients with COVID-19, which were presented in Figures 1 to 3. For pregnant samples, pre-pregnancy BMI was used. Data are shown as the mean ± SEM. *p < 0.05, **p < 0.01, ***p < 0.001.

**Figure S15.**
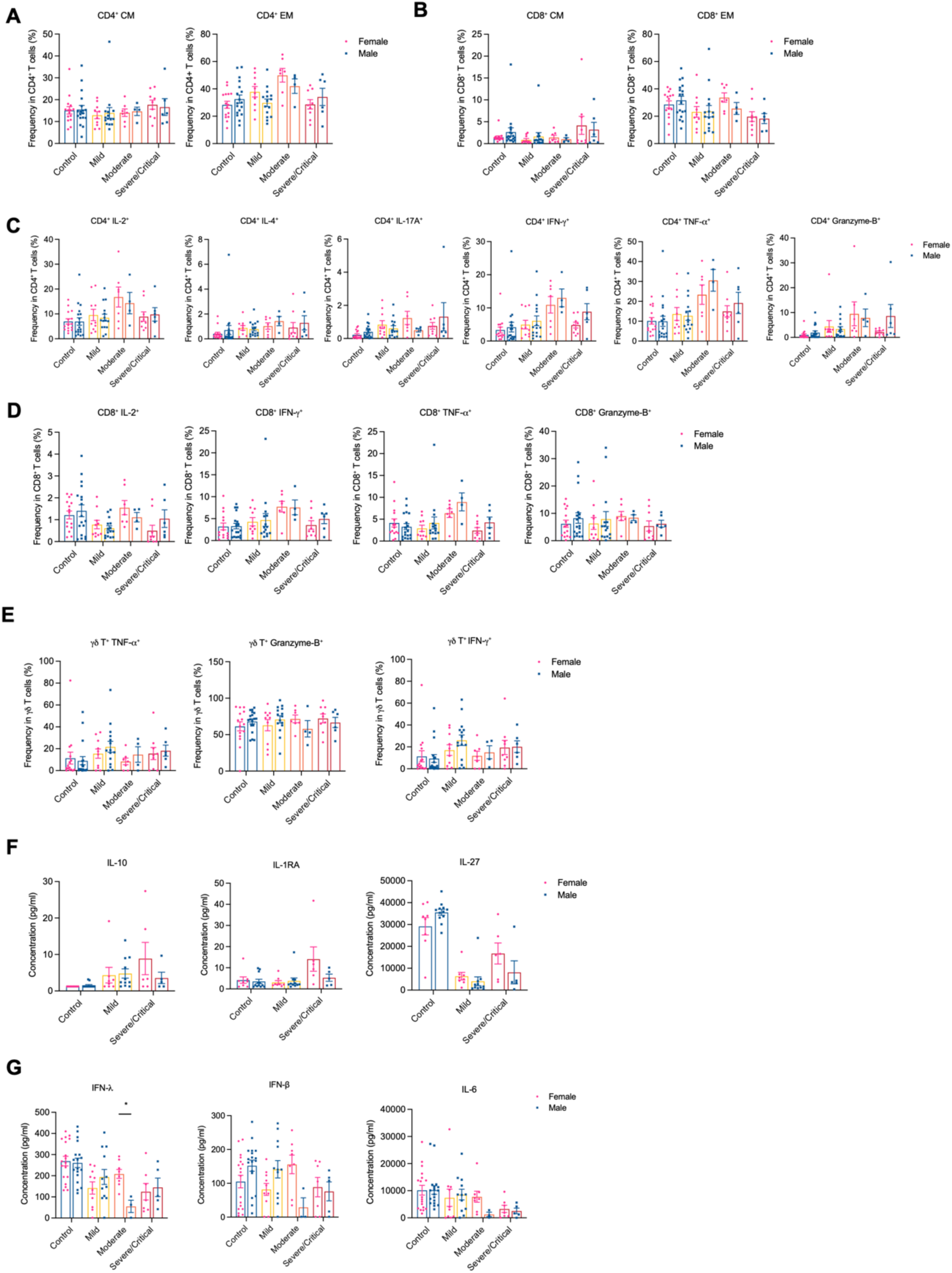
Effect of fetal sex on the immune responses of pregnant patients with COVID-19. (**A and B**) Naive, CM, EM, and EMRA frequencies in CD4^+^ T cells (**A**) and CD8^+^ T cells (**B**) were measured by flow cytometry. Healthy pregnant (female n = 15, male = 18) or who had mild (female n = 10, male = 15) or moderate (female n = 7, male = 4) or severe/critical (female n = 9, male = 6) in pregnant women. (**C** to **E**) Cytokine-producing CD4^+^ T cells (**C**), CD8^+^ T cells (**D**), and γδ-T cells (**E**) were measured by flow cytometry. Healthy pregnant (female n = 15, male = 18) or who had mild (female n = 10, male = 15) or moderate (female n = 7, male = 4) or severe/critical (female n = 9, male = 6) in pregnant women. (**F**) Plasma IL-10, IL-1RA, IL-27, and IFN-γ levels in control, mild, and severe/critical pregnant COVID-19 were measured. Healthy pregnant (female n = 8, male = 12) or who had mild (female n = 8, male = 11) or severe/critical (female n = 6, male = 5) in pregnant women. (**G**) Representative cytokines IFN-λ, IL-6, and IFNβ are presented as graphs in healthy pregnant (female n = 18, male = 18) or who had mild (female n = 10, male = 12) or moderate (female n = 8, male = 3) or severe/critical (female n = 7, male = 5) in pregnant women. Data are shown as the mean ± SEM. *p < 0.05.

**Figure S16.**
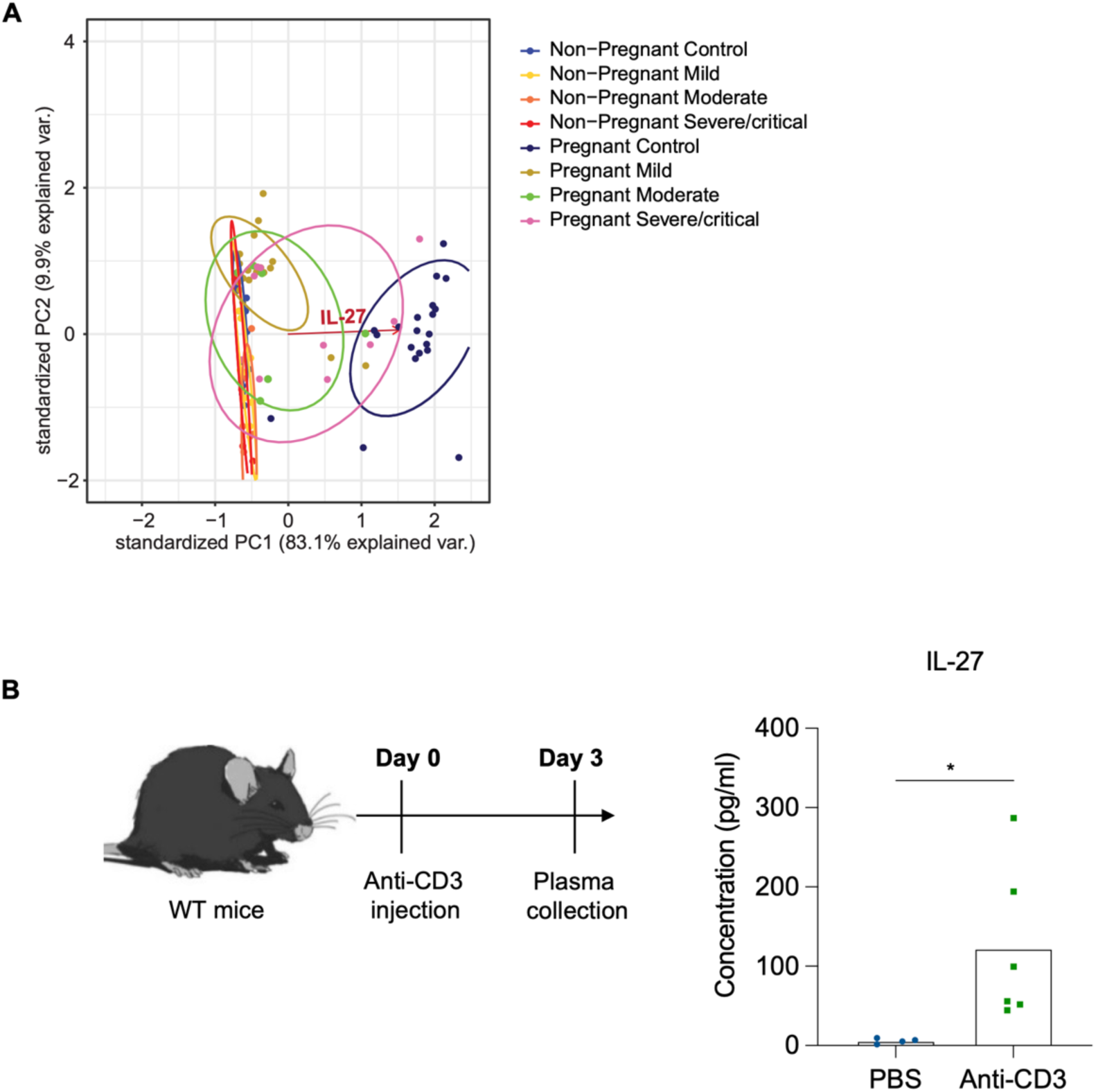
PCA analysis of plasma cytokine expression in pregnant COVID-19 and Anti-CD3 injection induces robust IL-27 expression. **(A)** PCA was applied to plasma cytokine expression for non-pregnant COVID-19 and pregnant COVID-19 subjects. The X and Y axes show principal components and percent variation of components 1 and 2, respectively. **(B)** WT mice were intraperitoneally injected with 20 μg of anti-CD3. ELISA was used to measure three days post-injection plasma IL-27 level. *p < 0.05.

**Tables** (**Table S3 to S10** were uploaded separately)

| Total participant women number | Non-pregnant | Pregnant |
| --- | --- | --- |
| Healthy control | 19 | 56 |
| Asymptomatic COVID-19 | 6 | 3 |
| Mild COVID-19 | 6 | 37 |
| Moderate COVID-19 | 5 | 14 |
| Severe/critical COVID-19 | 20 | 17 |
| Convalescent COVID-19 | 18 | 25 |

**Table S1.** Total number of participants.

| PBMC-Cell population analysis | Non-pregnant | Pregnant |
| --- | --- | --- |
| Healthy control | 19 | 34 |
| Mild COVID-19 |  | 27 |
| Moderate COVID-19 |  | 12 |
| Severe/critical COVID-19 | 9 | 15 |
| Convalescent COVID-19 | 18 | 22 |
| PBMC-Intracellular cytokine staining | Non-pregnant | Pregnant |
| Healthy control | 19 | 34 |
| Mild COVID-19 |  | 27 |
| Moderate COVID-19 |  | 12 |
| Severe/critical COVID-19 | 9 | 15 |
| Convalescent COVID-19 | 18 | 22 |
| PBMC-Monocyte analysis | Non-pregnant | Pregnant |
| Healthy control | 19 | 37 |
| Mild COVID-19 |  | 24 |
| Moderate COVID-19 |  | 12 |
| Severe/critical COVID-19 | 8 | 12 |
| Convalescent COVID-19 | 14 | 21 |
| Plasma profiling | Non-pregnant | Pregnant |
| Healthy control | 17 | 21 |
| Mild COVID-19 case | 12 | 21 |
| Moderate COVID-19 case | 9 | 6 |
| Severe/critical COVID-19 case | 10 | 11 |
| scRNA sequencing | Non-pregnant | Pregnant |
| Healthy control |  | 3 |
| Asymptomatic COVID-19 | 6 | 3 |
| Severe/critical COVID-19 case | 11 | 3 |
| Stool metagenome sequencing | Non-pregnant | Pregnant |
| Healthy control |  | 29 |
| Mild COVID-19 case |  | 13 |
| Moderate COVID-19 case | 4 | 6 |
| Severe/critical COVID-19 case | 9 | 8 |

**Table S2.** The summary of human samples per experiment

**Table S3. Sample metadata:** Meta-data for all single-cell samples.

**Table S4. CD8T_clones:** A list of all the clones found in CD8 T cells.

**Table S5. ISGs:** A list of the ISGs used to define the ISG score shown in Figures 4B, D, F, and Figure S9C.

**Table S6 SARS-CoV2 specific epitopes:** A list of all the SARS-CoV-2 epitopes found to be identified by CD8 TCRs, shown in Figures 3F,G, as found by querying VDJdb.

**Table S7. VDJdb_hits_wAlleles:** A list of all epitopes found in VDJdb as putatively binding CD8 TCRs, including patient HLA allele.

**Table S8.** Differentially expressed genes between cell subsets across all lineages.

**Table S9.** TotalSeqC antibodies that were used for CITEseq and hashtags used for hashing.

**Table S10.** Compilation of software packages used for the single cell data analysis.

